# Clinical outcomes associated with Omicron (B.1.1.529) variant and BA.1/BA.1.1 or BA.2 subvariant infection in southern California

**DOI:** 10.1101/2022.01.11.22269045

**Authors:** Joseph A. Lewnard, Vennis X. Hong, Manish M. Patel, Rebecca Kahn, Marc Lipsitch, Sara Y. Tartof

## Abstract

The Omicron (B.1.1.529) variant of SARS-CoV-2 rapidly achieved global dissemination following its emergence in southern Africa in November, 2021.^1,2^ Epidemiologic surveillance has revealed changes in COVID-19 case-to-hospitalization and case-to-mortality ratios following Omicron variant emergence,^3–6^ although interpretation of these changes presents challenges due to differential protection against Omicron or Delta (B.1.617.2) variant SARS-CoV-2 infections associated with prior vaccine-derived and naturally-acquired immunity, as well as longer-term changes in testing and healthcare practices.^7^ Here we report clinical outcomes among 222,688 cases with Omicron variant infections and 23,305 time-matched cases with Delta variant infections within the Kaiser Permanente Southern California healthcare system, who were followed longitudinally following positive outpatient tests between 15 December, 2021 and 17 January, 2022, when Omicron cases were almost exclusively BA.1 or its sublineages. Adjusted hazard ratios of progression to any hospital admission, symptomatic hospital admission, intensive care unit admission, mechanical ventilation, and death were 0.59 (95% confidence interval: 0.51-0.69), 0.59 (0.51-0.68), 0.50 (0.29-0.87), 0.36 (0.18-0.72), and 0.21 (0.10-0.44) respectively, for cases with Omicron versus Delta variant infections. In contrast, among 14,661 Omicron cases ascertained by outpatient testing between 3 February and 17 March, 2022, infection with the BA.2 or BA.1/BA.1.1 subvariants did not show evidence of differential risk of severe outcomes. Lower risk of severe clinical outcomes among cases with Omicron variant infection merits consideration in planning of healthcare capacity needs amid establishment of the Omicron variant as the dominant circulating SARS-CoV-2 lineage globally, and should inform the interpretation of both case- and hospital-based surveillance data.

---

Following its first detection in Gauteng Province, South Africa, the Omicron (B.1.1.529) variant of severe acute respiratory syndrome coronavirus 2 (SARS-CoV-2) was declared by the World Health Organization (WHO) to be a variant of concern on November 25, 2021.^1^ Rapid transmission of the Omicron variant fueled a fourth wave of SARS-CoV-2 infections in South Africa, during which daily diagnosed infections soon exceeded totals recorded during all previous periods in the country. Following its initial detection in the United States on 1 December, 2021,^8^ the Omicron variant rapidly became the dominant circulating lineage, accounting for 95% of all SARS-CoV-2 infections diagnosed nationwide by the week ending January 8, 2022.^9^ Similar patterns have unfolded globally, with the Omicron variant fueling a surge in newly-diagnosed cases worldwide.^10^ Across the US, prevalence of infection-derived antibodies increased from 34% to 58% during the Omicron wave between December, 2021 and February, 2022, and from 44% to 75% among children aged 0-11 years.^11^ While BA.2-lineage Omicron infections have subsequently accounted for increased transmission in March and April, 2022, increases in hospital admissions and deaths have not been commensurate with prior surges.^12^

Understanding the clinical spectrum of infections associated with novel SARS-CoV-2 variants is crucial to informing public health responses. Questions about the severity of Omicron variant infections arose soon after its emergence, as the Omicron genome harbored a constellation of mutations in the SARS-CoV-2 spike protein associated with altered cell entry as well as immune evasion.^13^ Reduced neutralization of the Omicron variant has been reported in studies using plasma specimens from individuals with complete (two- or three-dose) mRNA vaccine series,^14^ and from patients with prior SARS-CoV-2 infection.^15,16^ Epidemiologic data from South Africa have suggested higher rates of Omicron variant infections among persons with prior SARS-CoV-2 infection, as compared to observations with previous variants,^17^ while early observational studies in multiple settings have suggested reduced effectiveness of COVID-19 vaccines against Omicron variant infection.^18–20^ Notwithstanding these signs of reduced immune protection against the Omicron variant associated with prior natural infection or vaccination, increases in SARS-CoV-2 infections following emergence of the Omicron variant were not associated with increases in hospitalizations and deaths to the extent observed during previous waves.^3–6^ While reduced risk of hospitalization, intensive care unit (ICU) admission, and death has been reported among individuals with Omicron variant infection in several large-scale studies linking case data across various nationwide surveillance platforms,^20–22^ these studies have lacked detailed information about individual-level risk factors that may confound the relationship between infecting variant and risk of severe clinical outcomes. Understanding of the relative severity of disease associated with the BA.2 Omicron subvariant, which has become established in transmission despite widespread immunity from the initial Omicron wave, remains limited as well.^23,24^

## Study setting and variant dynamics

We sought to compare clinical outcomes among cases with Omicron and Delta variant SARS-CoV-2 infections within the Kaiser Permanente of Southern California (KPSC) healthcare system. As an integrated healthcare organization serving 4.7 million individuals (∼19% of the population of southern California), KPSC provides comprehensive care to its members across virtual, outpatient, emergency department, and inpatient settings. Healthcare delivery including diagnoses, laboratory tests and results, and prescriptions are recorded in near real-time via patients’ electronic health records (EHRs), while out-of-network care is captured through insurance claim reimbursements, enabling near-complete ascertainment of healthcare interactions for KPSC members.

Our primary analyses included all cases first ascertained via outpatient SARS-CoV-2 reverse transcription-polymerase chain reaction (RT-PCR) testing between 15 December, 2021 and 17 January, 2022, whose tests were processed using the ThermoFisher TaqPath COVID-19 Combo Kit (**Figure 1**; details on testing procedures are provided in the **Methods**). These dates comprise a period of mixed circulation of the two variants, with Omicron accounting for 99% of incident cases in the state of California by 17 January, 2022. Previous evidence has indicated that the Δ69-70 amino acid deletion in the spike (S) protein of Omicron variant specimens causes a failure in PCR probes targeting the S gene, whereas the Orf1ab and nucleocapsid (N) probes retain sensitivity; in contrast, S-gene target failure (SGTF) is rare in Delta variant SARS-CoV-2 infections (**Table S1**);^22,25,26^ thus, we used SGTF as a proxy for Omicron vs. Delta variant identification. Delta variant detections receded in late January as BA.2 Omicron subvariant infections began to account for an increasing proportion of all cases detected through February and March. We therefore also sought to investigate differences in risk of severe clinical outcomes among outpatient-detected cases BA.2 vs. BA.1* Omicron subvariant infections over the period of 3 February to 17 March, 2022, when reductions in Delta variant detection to <0.1% of incident cases made S gene detection a reliable proxy for BA.2 subvariant determination, consistent with observations in other settings.^23,24^

**Figure 1:**
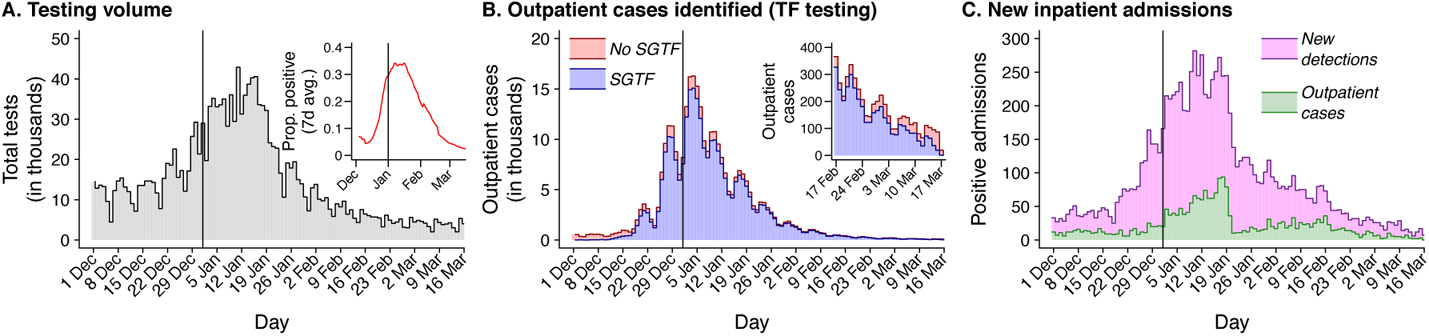
SARS-CoV-2 infections during follow-up within the study cohort. Plots illustrate total SARS-CoV-2 testing undertaken within the KPSC healthcare system across all clinical settings (**a**, along with the proportion of tests with positive results [*inset*]); total outpatient SARS-CoV-2 testing implemented using the ThermoFisher TaqPath COVID-19 Combo Kit assay along with the proportion of tests with SGTF identified (**b**; blue for SGTF detections and red for non-SGTF detections, with cases from 17 February to 17 March presented on an expanded scale for clarity [*inset*]); and new inpatient admissions of cases with SARS-CoV-2 infection (**c**; pink for new detections on or after the admission date and green for cases first ascertained by outpatient testing). Plotted data include 382,971 cases diagnosed over the study period, including 375,642 were tested in outpatient settings and 316,785 had samples processed using the ThermoFisher TaqPath COVID-19 Combo Kit assay.

## Characteristics of cases, by infecting variant

From 15 December, 2021 to 17 January, 2022, outpatient-diagnosed cases with Omicron variant infection (*N*=222,688) were concentrated among adults aged 20-39 years, and had lower odds of being either very young or very old in comparison to contemporaneously-identified cases with Delta variant infection (*N*=23,305; **Table S2**; **Table S3**). Cases with Omicron variant infection were also more often white and of non-Hispanic ethnicity, lived in higher-income communities, and tended to have lower prior-year rates of healthcare utilization across outpatient, emergency department, and inpatient settings, as well as lower burden of chronic comorbid conditions, in comparison to Delta variant cases (**Table S2**). These associations held in analyses adjusting for all measured demographic and clinical attributes of cases, which further included sex, current/former cigarette smoking, body mass index, and documented prior SARS-CoV-2 infection and COVID-19 vaccination. Adjusted odds of a prior documented SARS-CoV-2 infection ≥90 days before cases first tested positive during the study period were 1.75 (1.39-2.19) fold higher among cases with Omicron variant infection than among cases with Delta variant infection. Additionally, prior receipt of vaccine series associated with greater degrees of immune protection were more common among cases with Omicron variant infection; for instance, adjusted odds of prior receipt of a single mRNA vaccine dose (B162b2/mRna-1273) and a single Ad.26.COV2.S dose were 1.38 (1.27-1.51) and 1.56 (1.44-1.70) fold higher among cases with Omicron as compared to Delta variant infection, while adjusted odds of receipt of ≥3 mRNA vaccine doses were 2.60 (2.47-2.75) fold higher among cases with Omicron variant infection (**Table S2**; **Table S4**).

From 3 February to 17 March, 2022, among individuals tested as outpatients, BA.2 Omicron subvariant cases (*N*=1,905) did not tend to differ from BA.1* subvariant cases (*N*=12,756) in demographic or clinical attributes, with the exception that BA.1* detection was more concentrated among cases aged 20-49-years than BA.2, which was comparatively more common among both children and older adults; additionally, cases with BA.2 subvariant infection had higher rates of prior-year emergency department utilization than cases infected with BA.1* lineages (**Table S5**). Consistent differences in anti-SARS-CoV-2 immunity among cases with BA.2 or BA.1* infection—based on number or timing of vaccine doses received, or documented history of infection—were not apparent (**Table S6**).

## Risk of severe outcomes associated with infecting variant

Following an outpatient diagnosis, cumulative 30-day risks of hospital admission, symptomatic hospital admission (defined as new inpatient admission ≤14 days after new-onset acute respiratory symptoms), intensive care unit (ICU) admission, onset of mechanical ventilation, and death among cases with Delta variant infection were 10.3, 9.7, 1.1, 0.7, and 0.7 per 1000 cases, respectively, for cases testing positive between 15 December, 2021 and 17 January, 2022 (**Figure 2a-e**). For cases with Omicron variant infection over the same period of time, 30-day risks for the same outcomes were 4.5, 3.9, 0.2, 0.1, and 0.1 per 1000 cases. To understand whether these differences in risk could be explained by observed demographic, clinical, and immunological characteristics of cases with Delta and Omicron variant infection, we estimated adjusted hazard ratios (aHRs) for progression to each of these endpoints using Cox proportional hazards models, stratified on cases’ testing dates to further account for potential changes in clinical or testing practices over time (detailed in the **Methods**). Over the 30 days following an outpatient diagnosis, aHRs comparing progression to any hospital admission and symptomatic hospital admission among Omicron vs. Delta variant cases were 0.59 (0.51-0.69) and 0.59 (0.51-0.68), respectively (**Table S7**). These estimates should be interpreted as a weighted average of instantaneous aHRs comparing cases testing positive for Omicron and Delta variant infections on the same date, over their full follow-up period.^27^ For higher-acuity endpoints of ICU admission, mechanical ventilation, and mortality, aHR estimates comparing Omicron to Delta variant cases over the 60 days following outpatient detection were 0.50 (0.29-0.87), 0.36 (0.18-0.72), and 0.21 (0.10-0.44), respectively.

**Figure 2:**
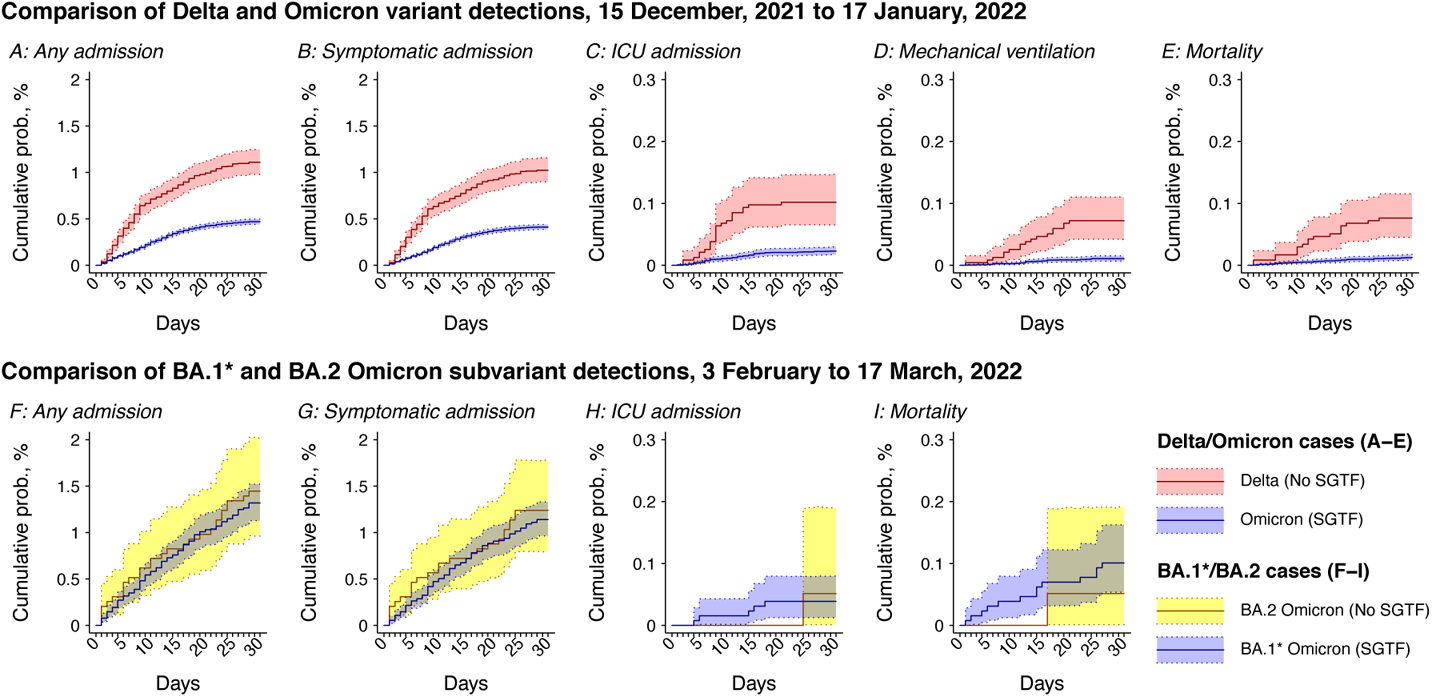
Severe clinical outcomes among cases. Plots illustrate cumulative 30-day risk of severe clinical outcomes among cases first ascertained in outpatient settings, stratified by SGTF status for infecting variant or subvariant. Panels in the top row compare cases with Delta (non-SGTF; red) or Omicron (SGTF; blue) variant infections testing positive in an outpatient setting between 15 December, 2021 and 17 January, 2022, for endpoints of any hospital admission (**a**); symptomatic hospital admission (**b**); intensive care unit admission (**c**); mechanical ventilation (**d**), and death (**e**). Panels in the bottom row compare cases with BA.2 (non-SGTF; yellow) and BA.1* (SGTF; blue, comprising BA.1/BA.1.1/BA.1.1.529 lineages) subvariant Omicron infections diagnosed in an outpatient setting between 3 February and 17 March, 2022, for endpoints of any hospital admission (**f**); symptomatic hospital admission (**g**); intensive care unit admission (**h**); and death (**i**). Mechanical ventilation among BA.2 and BA1* Omicron subvariant cases is not included due to sparse observations. Shaded areas denote 95% confidence intervals around point estimates. Analyses include 23,305 cases with Delta variant infection and 222,688 cases with Omicron variant infection over the period of 15 December, 2021 to 17 January, 2022, and 1,905 cases with BA.2 Omicron subvariant infection and 12,756 cases with BA.1* Omicron subvariant infection over the period of 3 February to 17 March, 2022. Confidence intervals are obtained via bootstrap resampling.

Similar estimates held in analyses that included cases diagnosed on or after their hospital admission date (**Table S7**), and in analyses restricted to cases who were asymptomatic at the point of testing (**Table S8**), among whom Omicron variant infection was also associated with modestly lower risk of subsequent symptoms onset (aHR=0.88 [0.81-0.96] for cases with Omicron vs. Delta variant infection tested in outpatient settings, without symptoms at the point of testing). Estimates of the aHR were also consistent in analyses restricted to cases with either complete data on measured covariates or those enrolled in KPSC health plans ≥1 year before their positive test date (**Table S9**); moreover, our findings held within subgroups defined on the basis of cases’ age, sex, presence of comorbidities, and history of documented SARS-CoV-2 infection (**Table S10**). Estimates of the adjusted relative risk (aRR) of hospital admission and symptomatic hospital admission (30-day) as well as ICU admission, mechanical ventilation, and mortality (60-day) based on log-binomial regression closely resembled aHR estimates from Cox proportional hazards models in the primary analysis (aRR=0.63 [0.55-0.72], 0.63 [0.55-0.72], 0.54 [0.31-0.94], 0.35 [0.17-0.71], 0.20 [0.10-0.43] for the five endpoints, respectively; **Table S11**). Findings of reduced risk of progression to hospital admission and symptomatic hospital admission further held within sensitivity analyses that additionally accounted for the possibility of differential prevalence of undiagnosed prior SARS-CoV-2 infection among cases with Omicron or Delta variant infection, who were or were not hospitalized, and who were or were not vaccinated (**Figure S1**; **Figure S2**).

We did not identify evidence of differences in risk of severe outcomes associated with BA.2 or BA.1* Omicron subvariant infection among cases diagnosed in outpatient settings over the period of 3 February to 17 March, 2022 (**Table S12**). Among cases with BA.1* Omicron subvariant infection diagnosed over this period, 30-day risks of hospital admission, symptomatic hospital admission, ICU admission, mechanical ventilation, and mortality were 13.3, 11.5, 0.4, 0.0, and 1.0 per 1000 persons, respectively (**Figure 2f-j**). Among cases with BA.2 infection, 30-day risks of the same outcomes were 14.7, 12.6, 0.5, 0.5, and 0.5, respectively per 1000 persons.

## Comparative severity of Delta and Omicron variant infections in subgroups by vaccination status

Coefficient estimates from Cox proportional hazards models suggested equivalent numbers of vaccine doses were associated with greater reductions in risk of severe outcomes among cases with Delta variant infection as compared to Omicron variant infection (for 2 mRNA doses ≤90 days prior to testing vs. 0 doses, aHR=0.17 [0.12-0.24] among Delta variant cases and aHR=0.51 [0.34-0.76] among Omicron variant cases; for 3 mRNA doses vs. 0 doses, aHR=0.14 [0.08-0.24] among Delta variant cases and aHR=0.43 [0.35-0.52] among Omicron variant cases; **Table S13**). Because variant-specific differences in vaccine protection could thus confound the relationship between infecting variant and risk of severe clinical outcomes, we further undertook analyses of cases with Delta and Omicron variant infection stratifying by prior vaccine exposure. For endpoints of hospital admission, symptomatic hospital admission, ICU admission, mechanical ventilation, and mortality, aHR estimates were 0.40 (0.33-0.49), 0.40 (0.33-0.49), 0.34 (0.17-0.66), 0.24 (0.12-0.48), and 0.14 (0.07-0.28), respectively, among unvaccinated cases with Omicron versus Delta variant infection (**Figure 3**; **Table S14**). In contrast, variant-specific differences in risk of hospital admission or symptomatic hospital admission were not apparent among individuals who received ≥3 mRNA vaccine doses. Among individuals who had received 2 mRNA vaccine doses, differences in risk of hospital admission or symptomatic hospital admission were likewise attenuated, with the smallest difference among individuals most recently vaccinated. We did not identify differences in risk of ICU admission and mechanical ventilation among vaccinated cases with Omicron or Delta variant infection. However, among vaccinated individuals, Omicron infection remained associated with a lower risk of mortality than Delta infection (aHR=0.25 [0.09-0.70]). Similar findings held in analyses that included individuals testing positive on the day of hospital admission (**Figure S3**).

**Figure 3:**
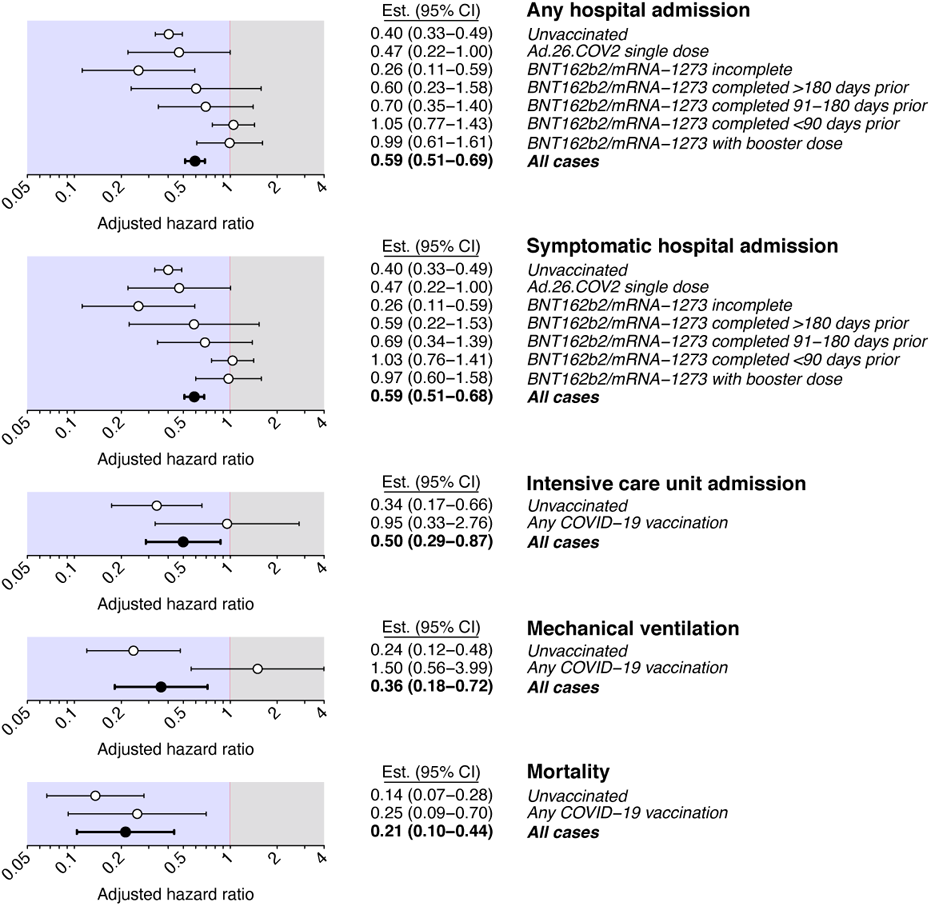
Adjusted hazard ratios of severe clinical endpoints within strata defined by vaccination status. Points and lines denote estimates and accompanying 95% confidence intervals for the adjusted hazard ratio of each endpoint, comparing cases with Omicron versus Delta variant infection, in case strata defined by history of COVID-19 vaccination. Analyses are restricted to individuals tested diagnosed in outpatient settings by RT-PCR testing using the ThermoFisher TaqPath COVID-19 combo kit; adjusted hazard ratios are estimated using Cox proportional hazards regression models, controlling for covariates listed in **Table S2** and stratifying on positive test date. Analyses include 23,305 cases with Delta variant infection and 222,688 cases with Omicron variant infection. Confidence intervals are obtained using Cox proportional hazards regression models.

## Comparing risk of severe clinical outcomes during periods of Delta and Omicron variant predominance

Because excluding cases whose tests were not processed using the ThermoFisher TaqPath COVID-19 combo kit could limit the generalizability of our primary analyses, we also assessed changes over the period of 1 November, 2021 (prior to detection of the Omicron variant in the state of California) to 17 January, 2022 in the risk for all cases diagnosed in outpatient settings to progress to severe clinical outcomes. In analyses using Cox proportional hazards models which allowed for zero, one, or two changepoints in the relationship between testing date and risk of severe clinical outcomes,^28^ we identified evidence for a reduction beginning 8-23 December, 2021, in cases’ risk of any hospital admission, symptomatic hospital admission, intensive care unit admission, and mortality among newly-diagnosed cases (**Figure 4a-e**; **Table S15**; changepoint models were not fitted to the mechanical ventilation endpoint due to sparse observations during the Delta variant-dominated period). This timing encompasses the period of Omicron’s expansion in the study population, with 5% and 50% of cases tested on ThermoFisher TaqPath COVID-19 combo kit assays showing SGTF as of 10 and 17 December, 2021, respectively.

**Figure 4:**
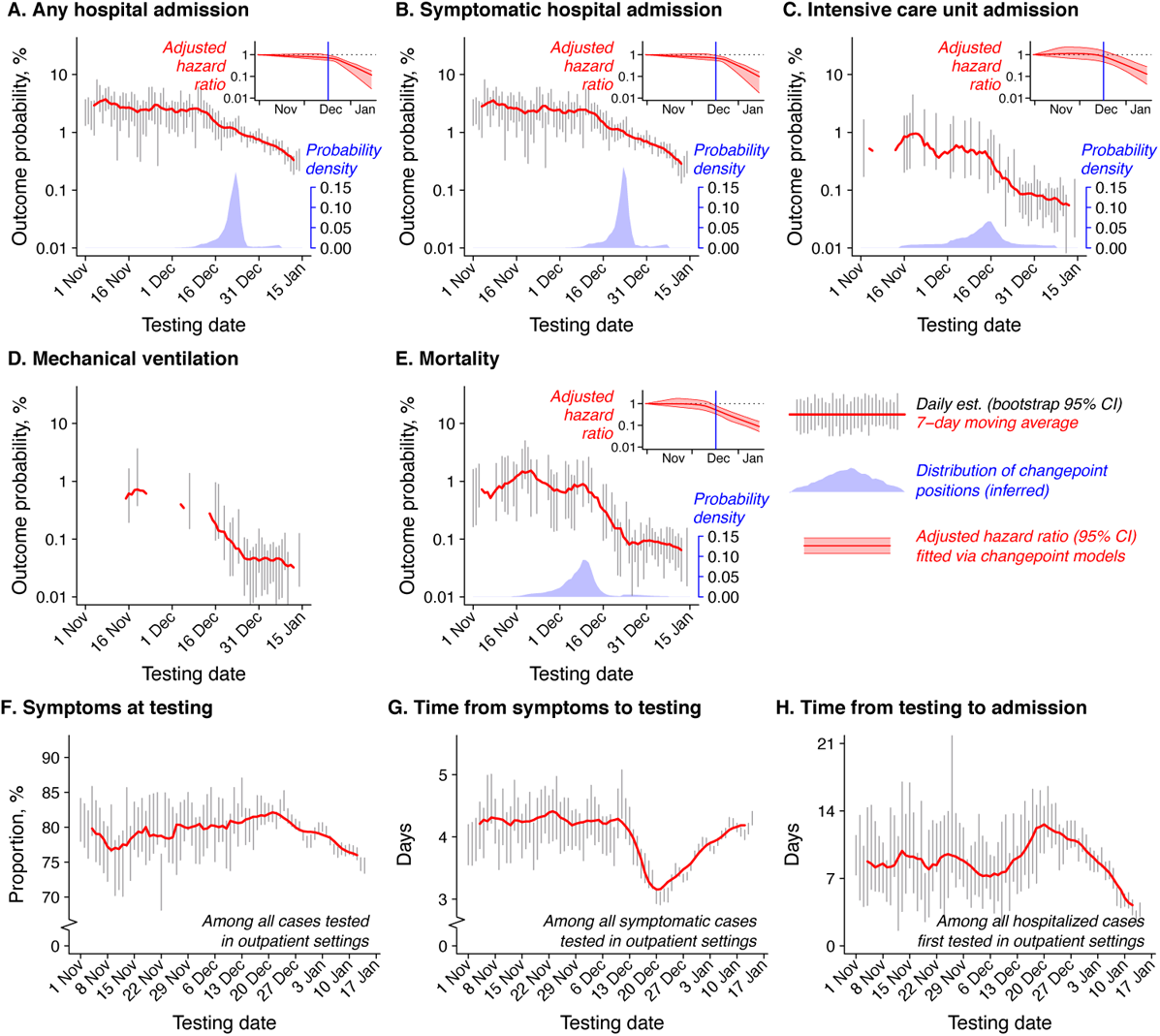
Changes in risk of severe clinical outcomes and in symptoms history among cases during the study period. Panels illustrate proportions of cases experiencing each clinical outcome over the course of follow-up (30 days for endpoints of hospital admission [**a**] or symptomatic hospital admission [**b**]; 60 days for ICU admission [**c**], mechanical ventilation [**d**], and mortality [**e**]). Gray lines denote 95% confidence intervals around estimates for each day based on bootstrap resampling; 7-day moving averages are plotted in red lines. Polygons at the bottom of panels **a**-**e** illustrate probability densities of change point timings (blue), while inset panels illustrate fitted slopes for adjusted hazard ratio (aHR) estimates for each endpoint as a function of testing date (red; lines indicating point estimates and polygons delineating 95% confidence intervals). Bottom panels illustrate the proportion of cases tested in outpatient settings indicating symptoms onset on or before their testing date (**f**); mean time from symptoms onset to outpatient testing, among symptomatic cases (**g**); and mean time from the testing date to hospital admission, among admitted cases (**h**). Changes in the proportion of cases ascertained in inpatient settings are plotted separately in **Figure S4**. Analyses include 316,038 outpatient-diagnosed cases. Confidence intervals around proportions of cases experiencing severe outcomes, by testing date, are obtained by bootstrap resampling; confidence intervals in inset plots around the adjusted hazard ratio of severe outcomes are obtained using Cox proportional hazards regression models.

Observed reductions in cases’ risk of severe outcomes did not directly align with changes in clinical attributes of cases testing positive in outpatient settings over this period, suggesting that shifting patterns of healthcare utilization and clinical practice could not fully account for the observed changes. The proportion of cases reporting symptoms on or before their testing date held steadily in the range of 72.2-84.3% from 1 November, 2021 to 17 January, 2022 (**Figure 4f**). While the mean time from symptoms onset to testing (among symptomatic cases) dipped transiently to 3.2 days between December 19-22 (as compared to 4.2 days in November and mid-January; **Figure 4g**), time from testing to inpatient admission (among cases ultimately requiring hospitalization) declined through the month of January, consistent with cases seeking outpatient testing at a more advanced stage of their illness (**Figure 4h**). Concurrently, the proportion of all SARS-CoV-2 infections detected in inpatient settings declined from 2.4% to 0.7% between 1 and 31 December, 2021, although this proportion increased roughly 10-fold to 7.8% as of 7 March, 2022 amid reductions in outpatient testing volume during 2022 (**Figure S4**).

## Lengths of hospital stay

Duration of hospital stay among cases whose illness meets the severity threshold for hospital admission provides additional insight into differences in the clinical course of SARS-CoV-2 variants.^29,30^ Among 208 cases testing positive for Delta-variant infection in outpatient settings and admitted to hospital over the period of December 15, 2021 to 17 January, 2022, the proportions with hospital stays lasting ≤5 days, ≤10 days, and ≤15 days were 66.2%, 84.5%, and 89.4%, respectively, in comparison to 84.8%, 91.0%, and 92.2% among 703 cases with Omicron variant infection tested and admitted over the same period (**Figure 5a-f**; **Table S16**). Within this sample, 73.8% and 85.6% of admitted cases with Delta and Omicron variant infections, respectively, were discharged home within ≤30 days, while 15.5% and 6.0% of admitted cases with Delta and Omicron variant infections, respectively, were referred to other care settings or discharged against medical advice within the same timeframe. The 30-day probability of death or discharge to hospice following admission was 1.1% for cases with Delta variant infection and 0.4% for cases with Omicron variant infection. Using a Cox proportional hazards model stratified on cases’ admission date and controlling for all observed demographic, clinical, and immunological attributes of cases to compare time to completion of hospital stay (with any final disposition), the aHR comparing outpatient-diagnosed cases with Omicron vs. Delta variant infection was 1.24 (0.99-1.57; **Table S17**). No differences in the duration of hospital stay, or likelihoods of each discharge disposition, were evident among outpatient-diagnosed cases with BA.2 or BA.1* Omicron subvariant infection tested and admitted between 3 February and 17 March, 2022 (**Figure 5g-k**). The aHR for completion of hospital stay (with any final disposition) for outpatient-diagnosed cases with BA.2 vs. BA.1* Omicron subvariant infection was 0.95 (0.41-2.22; **Table S17**).

**Figure 5:**
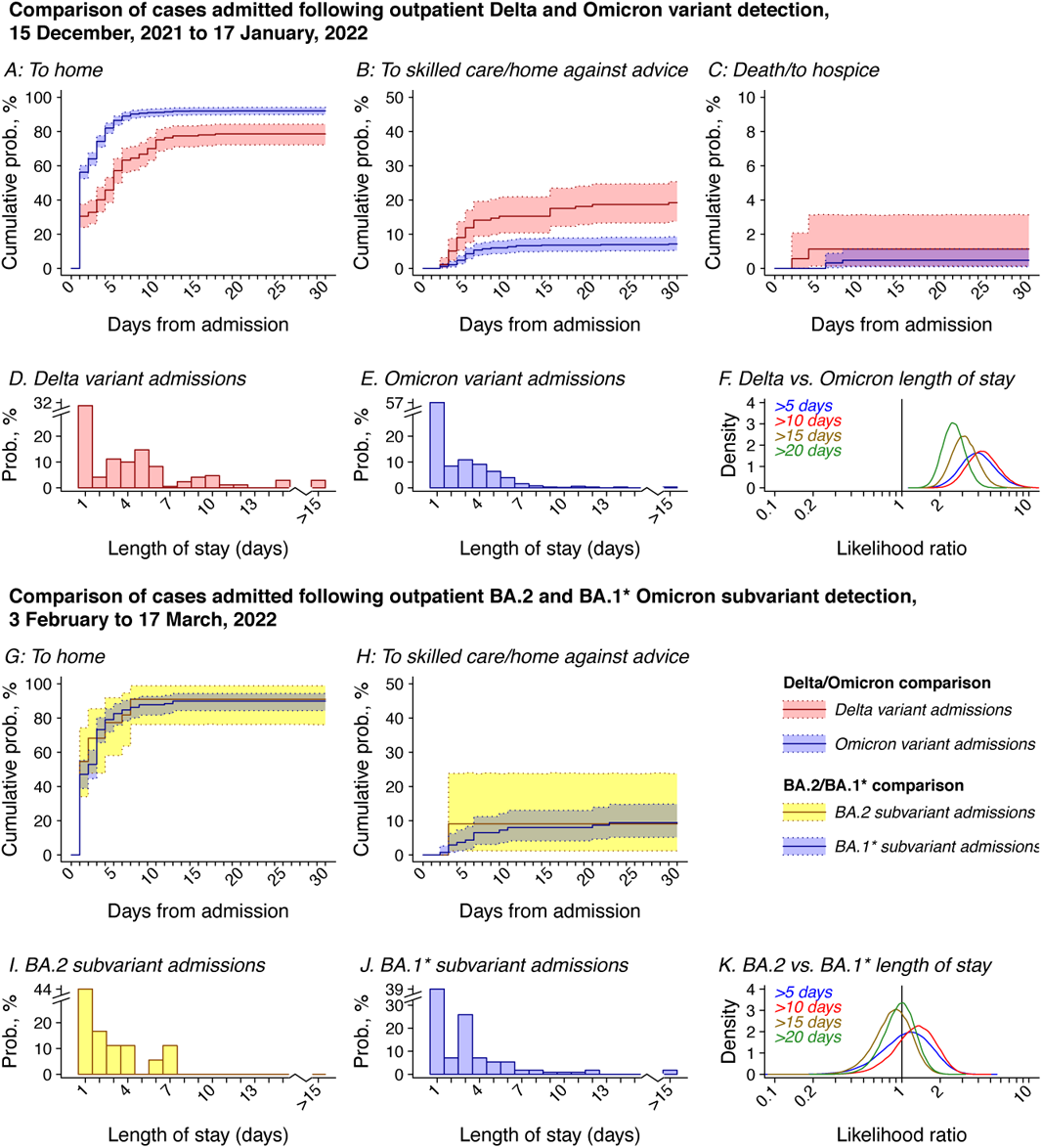
Durations of hospital stay. Top panels illustrate times from hospital admission to discharge to home without skilled care (**a**), discharge to skilled care or against medical advice (**b**), and in-hospital death or discharge to hospice (**c**) among cases testing positive in outpatient settings and subsequently admitted to hospital on or after the date of symptoms onset over the period from 15 December, 2021 to 17 January, 2022; lines and polygons indicate point estimates and 95% confidence intervals, respectively, based on bootstrap resampling for cases with Delta (red) and Omicron (blue) variant infection. Below, panels illustrate histograms of the total length of stay for cases with Delta variant infection (**d**) and Omicron variant infection (**e**) within this sample, as well as distributions of the likelihood ratio for cases with Delta vs. Omicron infection to have hospital stays lasting >5 days, >10 days, >15 days, and >20 days (**f**). The bottom set of panels illustrates times from admission to discharge to home (**g**), and discharge to skilled care or against medical advice (**h**), among cases testing positive in outpatient settings and subsequently admitted to hospital on or after the date of symptoms onset over the period from 3 February to 17 March, 2022; lines and polygons indicate point estimates and 95% confidence intervals, respectively, based on bootstrap resampling for cases with BA.2 (yellow) and BA.1* (blue) Omicron subvariant infection. Below, panels illustrate histograms of the total length of stay for cases with BA.2 Omicron subvariant infection (**i**) and BA.1* Omicron subvariant infection (**j**) within this sample, as well as distributions of the likelihood ratio for cases with BA.2 vs. BA.1* Omicron subvariant infection to have hospital stays lasting >5 days, >10 days, >15 days, and >20 days (**k**). Analyses include 208 cases with Delta variant infection and 703 cases with Omicron variant infection for the period of 15 December, 2021 to 17 January, 2022, and 23 cases with BA.2 Omicron subvariant infection and 146 cases with BA.1* Omicron subvariant infection for the period of 3 February to 17 March, 2022. Confidence intervals are computed via bootstrap resampling.

## Discussion

Among cases followed from an outpatient SARS-CoV-2 diagnosis, infection with the Omicron variant was associated with substantially lower risk of progression to severe clinical outcomes including hospital admission, symptomatic hospital admission, ICU admission, mechanical ventilation, and mortality, in comparison to infection with the Delta variant. These differences in risk among individuals with Omicron versus Delta variant infection were consistent with reductions in the proportion of all SARS-CoV-2 cases that progressed to severe clinical outcomes during the period of Omicron variant emergence in the study population. We also observed shorter durations of hospital stay following inpatient admission among cases with Omicron as compared to Delta variant infection. Whereas admitted cases with Omicron variant infection had higher likelihood than cases with Delta variant infection of being discharged to home, those with Delta variant infection had higher probability of mortality and discharge to skilled care or against medical advice. We did not identify evidence of differences in severity for cases with BA.2 and BA.1* Omicron subvariant infection, based on either their risk of severe clinical outcomes or their hospital length of stay and final disposition following inpatient admission. Collectively, these findings suggest that cases with Omicron variant infections face reduced risk of severe clinical outcomes, and that this association has held following emergence and establishment of the BA.2 Omicron subvariant.

Previous studies have estimated reductions in risk of hospital admission associated with Omicron variant infection across a range spanning 20-80%. ^20–22,31–34^ Variability in prior estimates from database linkage studies may owe in part to intra- and inter-study differences in immunity, health status, and healthcare-seeking behaviors among cases across settings. As KPSC serves as a comprehensive healthcare provider to its members, and tracks out-of-network care provision for its members through insurance claim reimbursements, our study benefited from highly-resolved electronic health records as a basis for characterizing cases’ clinical history. Similar detail may be lacking in other large-scale studies from throughout the pandemic, which have varyingly relied on administrative record linkage to identify comorbid conditions,^21^ had access to such data only for admitted cases based on in-hospital assessment or record linkage,^22,32,35^ or have lacked data on cases’ history of comorbidities and healthcare utilization entirely.^20,34,36,37^ Despite these differences in specific design features and estimates across studies, consistency of the finding that Omicron variant infection is associated with reduced severity relative to Delta variant infection is noteworthy.

Several other aspects of our study helped to control for relevant potential differences in attributes of cases with Delta and Omicron variant infection, which could otherwise confound comparisons of risk for severe clinical outcomes. Stratification of Cox proportional hazards models on cases’ testing and admission dates, and inclusion of day-specific intercepts in logistic regression models, helped to correct for potential differences in attributes of cases tested or admitted over time unrelated to the infecting variant. Restriction of primary analyses to cases tested in outpatient settings enabled us to account for selection on healthcare-seeking behavior among cases infected with either variant. This strategy further standardized the level of clinical severity associated with the hospital admission endpoint. Following outpatient testing at KPSC throughout the study period, cases considered to be at risk of severe illness, but not meeting admission criteria, were enrolled in a home-based monitoring program with daily clinical interaction and standardized criteria for emergency department referral and inpatient admission.^38^ Thus, clinical severity at the point of admission among outpatient-diagnosed cases may have been less variable than among cases ascertained at the point of admission. Focusing primary analyses on outpatient-diagnosed cases also helped to limit inclusion of hospitalizations attributable to causes other than COVID-19. As the incidence rate of all-cause hospital admissions is low, few hospitalizations attributable to factors other than COVID-19 would be expected to occur within the short period of time immediately following a positive outpatient test; in contrast, a substantial proportion of SARS-CoV-2 infections among hospitalized cases may be detected simply due to entry screening at the point of admission for other causes.^39^ While it is a limitation that no routinely-collected records provide a gold standard determination of whether SARS-CoV-2 infection was the cause of the decision to admit a patient to hospital, these factors (together with our consideration of one endpoint restricted to inpatient admissions occurring on or after the date of symptoms onset) limit the risk of misclassification of hospital admissions attributable to causes other than COVID-19 to a greater extent than has been possible in prior studies of Omicron as well as other SARS-CoV-2 variants.^4,17, 20–22,32,33,37,40^

Unobserved prior infections are a potential source of bias when comparing risk of severe outcomes among cases with Omicron or Delta variant infection.^41,42^ Prior to the Omicron epidemic wave, roughly one in 2.5 infections in the state of California were estimated to have been caught by testing, indicating cases’ history of infection may be substantially underestimated in our study population.^43^ Moreover, because convalescent sera from previously-infected individuals has shown weaker neutralization activity against the Omicron variant as compared to Delta (and earlier) variants,^44,45^ prevalence of unascertained prior infection among cases with Omicron and Delta variant infections may be differential. We nonetheless identified that findings of reduced severity of Omicron variant infections held within bias analyses allowing substantially greater-than-observed prevalence of prior infection among previously-vaccinated cases with Omicron variant infections who were not hospitalized—the stratum within which unobserved infections would contribute the greatest degree of bias for our analyses. In agreement with these findings, sensitivity analyses within prior studies using diverse statistical inference methods have suggested that differences in risk of severe clinical outcomes among cases with Omicron and Delta variant infections cannot be explained by unobserved prior infections alone.^17,20^ Furthermore, the unadjusted HR of hospital admission associated with Omicron variant detection among cases in our study known to have experienced prior infection was 0.27 (0.03-2.44) following a positive outpatient test (**Table S10**); although analyses within this stratum are underpowered, the direction of association is telling as differential unobserved prior infection among cases with Omicron and Delta variant infection could not account for risk of severe outcomes among these cases.

There are several barriers and limitations to causal inference in this study. Because our analysis aims to compare disease severity among cases following acquisition of Omicron or Delta variant infection, no real-world trial directly emulates the inferential design conditioning on acquisition of infection. Observed associations of infecting lineage with case attributes within our sample should not be considered to represent predictors of acquisition of a specific infecting SARS-CoV-2 lineage;^46^ risk factors for exposure to each variant and for infection, given exposure, are outside the scope of this study.

While statistical adjustment for differences in demographic, clinical, and immunological aspects of cases supported efforts to define associations of each variant with risk of severe outcomes, given acquisition of infection, unobserved attributes of cases which predict both their infecting variant and risk of severe clinical outcomes remain of concern, as in all observational epidemiologic research.^47^ Last, while selecting cases who sought outpatient tests ensures our primary analysis encompasses individuals meeting a minimum threshold for healthcare-seeking behavior, further adjustment for this characteristic was limited to cases’ prior-year frequency of healthcare utilization across outpatient, emergency department, and inpatient settings. Notwithstanding these limitations, our findings of reduced severity in Omicron variant infections are consistent with numerous lines of experimental evidence not susceptible to similar sources of bias. *Ex vivo* studies demonstrate higher replication of the Omicron variant in the human upper respiratory tract as compared to the small airways of the lung,^48^ while in animal models disease associated with the Omicron variant has be confined to the large airway.^49^

While attenuation of disease severity in Omicron variant infections—which has held amid emergence of the BA.2 subvariant—is an encouraging finding, evidence of higher transmissibility of Omicron variant infections^50^ as well as immune evasion from prior infection and vaccination remain concerning. High rates of infection in the community have overwhelmed health-care systems within the US and other settings, and have translated to high absolute numbers of hospitalizations and deaths even with lower severity of infections associated with the Omicron variant. Observations in settings with low prevalence of infection-derived immunity, such as Hong Kong^51^ and New Zealand,^52^ underscore the risk for the Omicron variant to cause substantial burden of severe and fatal illness even if cases tend to experience lower risk of severe clinical outcomes than with Delta variant infection. This observation is also consistent with the frequent occurrence of severe disease cases and deaths in settings such as long-term care facilities in the US, United Kingdom, and Italy with ancestral (Wuhan) variant infections.^53^ Our findings underscore the value of monitoring variant-specific infection severity alongside ongoing surveillance efforts aimed at tracking epidemiologic dynamics of novel variants to inform intervention deployment and healthcare capacity planning.

## Data Availability

Requests for access to data should be submitted to.

## METHODS

### Setting, procedures, and study population

Care delivery and EHR data capture in the KPSC healthcare system have been described previously.^54^ Briefly, members of KPSC receive care through employer-provided, pre-paid, or federally sponsored insurance plans and closely resemble the sociodemographic profile of the surrounding geographic area in terms of age, racial/ethnic composition, and community characteristics.^55^ Within-network care delivery encompassing diagnoses, laboratory tests and results, and prescriptions is captured in real time through patients’ EHR, while out-of-network care is captured through insurance reimbursements. COVID-19 vaccines were provided at no cost to KPSC members following emergency use authorization and were therefore captured in the EHR. Vaccinations administered outside KPSC were captured via the California Immunization Registry, to which providers are required to report all COVID-19 vaccine administrations within 24 hours. The study protocol was approved by the KPSC Institutional Review Board.

Polymerase chain reaction (PCR) testing for SARS-CoV-2 occurred in a variety of clinical settings within KPSC during the study period. A majority of tests conducted in outpatient settings are submitted to regional laboratories, where >90% of samples are processed using the ThermoFisher TaqPath COVID-19 Combo Kit. Samples collected in hospitals (including some tests conducted in emergency department settings) are processed using in-house tests, without SGTF readout. In total, 329,195 of 389,896 (84.4%) cases detected in outpatient settings from 1 November, 2021 to 17 March, 2022 had samples processed using the ThermoFisher TaqPath COVID-19 Combo Kit. Attributes of outpatient cases processed using the ThermoFisher TaqPath COVID-19 Combo Kit or other assays are presented in **Table S18**. Analyses comparing cases with Delta and Omicron variant infection were restricted to cases testing positive between 15 December, 2021 and 17 January, 2022, encompassing the period during which both variants were detected at >1% prevalence statewide in California, and preceding emergence of the BA.2 Omicron subvariant as a likely cause of S gene detection.^56^ Analyses comparing cases with BA.2 and BA.1* Omicron subvariant infection were restricted to cases testing positive between 3 February and 17 March, 2022, following the emergence of BA.2 at ≥1% frequency among all cases (while Delta accounted for <0.1%) and yielding ≥45 days of follow-up for all cases before the final database lock.

To assess variant-specific differences in risk of progression to severe endpoints in a time-to-event framework, our primary analyses included all cases diagnosed in outpatient settings with a positive PCR test processed on a ThermoFisher TaqPath COVID-19 combo kit during the study period, who were continuously enrolled in KPSC health plans through the relevant follow-up periods (detailed below) or until their death, whichever was earlier. Restricting to outpatient-diagnosed cases aimed to address two potential sources of bias, including (1) selecting on healthcare-seeking behavior to mitigate confounding that may occur with individuals who deferred testing to more severe stages of illness; and (2) limiting the inclusion of hospital admissions where SARS-CoV-2 infection was detected incidentally, for instance through entry screening. Because hospital admission is a rare event, the number of admissions attributable to factors other than COVID-19 in the time immediately following a positive SARS-CoV-2 test was expected to be low. To ensure our analyses addressed newly-acquired SARS-CoV-2 infections and not PCR-positive detections of remote infections, we excluded individuals with a prior positive SARS-CoV-2 testing result within ≤90 days before their first eligible positive result during the study period.

### Outcome measures

As primary endpoints, we considered five markers of clinically severe illness: any hospital admission, hospital admission associated with new-onset acute respiratory symptoms, ICU admission, mechanical ventilation, and mortality. Hospital admissions were considered to be COVID-19-related if they occurred between 7 days before to 30 days after the date of each patient’s positive SARS-CoV-2 RT-PCR test; we included ICU admissions, mechanical ventilation events, and deaths occurring up to 60 days after the date of each positive test in the analysis (or up to 45 days after the positive test date for analyses of cases with BA.2 or BA.1* Omicron subvariant infection).

Symptomatic hospital admissions were those with acute respiratory infection symptoms beginning on or ≤14 days before the admission date; we ascertained presence of symptoms and dates of symptoms onset via natural language processing of open-text EHR fields including clinical notes and patient-provided questionnaire responses, which are submitted by all KPSC patients who seek SARS-CoV-2 testing regardless of test setting.^54^ We considered new-onset respiratory symptoms following a positive test as a secondary endpoint for further exploratory analyses among cases who were asymptomatic at the time of their original test.

Last, for a duration-of-hospital-stay analysis, we recorded dates of discharge and discharge disposition, in-hospital mortality, or censoring for all hospitalized patients. Analyses were restricted to cases who were tested and admitted to hospital during the periods of 15 December, 2021 to 17 January, 2022 (for comparisons of cases with Delta and Omicron variant infection) and 3 February to 17 March, 2022 (for comparisons of cases with BA.2 and BA.1* Omicron subvariant infection); inclusion of cases diagnosed within these two periods ensured ≥60 days and ≥45 days follow-up for all cases from the point of admission. Duration-of-stay analyses included all eligible outpatient-diagnosed cases from the primary analysis cohort, whose samples were processed using the ThermoFisher TaqPath COVID-19 Combo Kit, and who experienced acute new-symptoms onset respiratory symptoms on or before their admission date.

### Considerations for hospital admission

As routine data capture does not include a “gold standard” indication as to whether COVID-19 or another factor served as the primary cause of physicians’ decision to admit a patient, we caution that factors other than SARS-CoV-2 infection (or in conjunction with SARS-CoV-2 infection) may have contributed to hospital admission outcomes as well as ICU admission, use of mechanical ventilation, and death, including among individuals with new-onset respiratory symptoms before their admission date, consistent with prior COVID-19 studies using hospital admission endpoints.^4,17, 20–22,32,33,37,40^ However, several measures implemented by KPSC to preserve hospital capacity during the COVID-19 pandemic may have lessened the capture of incidental admission events among outpatient-diagnosed cases within our sample. Outpatient administration of remdesevir and monoclonal antibody therapies was prioritized so that access to treatment would not be grounds for admission. In addition, KPSC used a scoring system to standardize admission versus outpatient management decisions throughout the study period based on cases’ clinical history and comorbidities (electrolyte disorders, cardiac arrhythmia, neurological disorders, weight loss disorders, congestive heart failure, coagulopathy, diabetes), body mass index, vital signs (oxygen saturation, respiratory rate, systolic blood pressure, fever, and heart rate), age, and sex.^38^ Based on the resulting scores at the point of testing, cases were recommended for one of three levels of care provision. Patients not recommended for inpatient admission were either sent home with a telemedicine follow-up from their primary care provider (lowest-risk patients) or enrolled in a home-based monitoring program, for which patients were provided a medical-grade pulse oximeter and thermometer, and instructed to enter readings daily into a mobile application to alert physicians in the event of clinical deterioration. Standardized criteria were used for subsequent emergency room referral and hospital admission during subsequent follow-up.

### Case attributes

Recorded characteristics of cases included: age (defined for all analyses as bands of <1, 1-4, 5-9, 10-19, 20-29, 30-39, 40-49, 50-59, 60-69, 70-79, and ≥80 years), sex, race/ethnicity (white, black, Hispanic of any race, Asian/Pacific Islander, and other/mixed/unknown race), census tract-level median household income (defined on the log scale for all analyses); smoking status (current, former, or never smoker); body mass index (BMI; underweight, normal weight, overweight, and obese); Charlson comorbidity index (0, 1-2, 3-5, and ≥6); prior-year emergency department visits and inpatient admissions (each defined as 0, 1, 2, or ≥3 events); prior-year outpatient visits (0-4, 5-9, 10-14, 15-19, 20-29, or ≥30 events); documented prior SARS-CoV-2 infection; and history of COVID-19 vaccination (unvaccinated, Ad.26.COV2.S as one dose or with a booster; and 1, 2, or 3 mRNA vaccine doses, disaggregating 2-dose recipients by time since receipt of the second dose as ≤90 days, 91-180 days, or ≥181 days). We compared the distribution of these attributes among cases with Delta and Omicron variant infection, and among cases with BA.2 or BA.1* Omicron subvariant infection, using logistic regression models defining intercepts for cases’ testing date and multiple imputation for missing values (as described below in the description of the primary analysis).

### Association of SGTF with risk of severe clinical outcomes

Within the primary analysis population, we compared times from the first positive test to each outcome event among patients who tested positive for SARS-CoV-2 by RT-PCR, with and without SGTF. We censored observations at 30 days for hospital admission and symptomatic hospital admission, and at 60 days for ICU admission, mechanical ventilation, and death (or at 45 days for these endpoints among cases included in the BA.2/BA.1* analyses). We used Cox proportional hazards models to estimate the aHR for each endpoint associated with SGTF, adjusting for all available demographic and clinical covariates according to the definitions provided above. We defined strata for cases’ testing date to account for potential secular changes in testing and healthcare practices over the study period, noting that testing dates were jittered at random by 0, +1, or −1 days to preserve anonymity of protected health information; lengths of time to event or censoring were preserved for analysis integrity. We additionally fit models allowing for interactions of SGTF sample status with cases’ vaccination history to assess variation in the estimated association of SGTF with vaccination status, as described in the primary results above.

### Sensitivity analyses

We repeated analyses of the symptomatic hospital admission endpoint within subgroups defined by patient age, sex, Charlson comorbidity index, and history of documented SARS-CoV-2 infection and vaccination, controlling for all other risk factors via covariate adjustment. We also conducted secondary analyses including all patients whose tests were processed using the ThermoFisher TaqPath COVID-19 combo kit, regardless of diagnosis setting. In these analyses, times to events were recorded as 0.5 days for patients experiencing study endpoints on or before the date of their test. These sensitivity analyses aimed to address any bias that could result from exclusion of cases who progressed rapidly to clinically-severe illness; results are presented in **Table S7** and **Figure S3**. We also conducted subgroup analyses within the sample of cases who did not experience symptoms onset on or before their testing date. As a greater likelihood of symptoms among cases infected with either of the two variants could obscure differences in variant-associated clinical severity (i.e., selecting on a differential subset of cases with the otherwise less-severe variant), these analyses aimed to capture a broader spectrum of the clinical course by monitoring cases from a point preceding symptoms onset. Results are presented in **Table S8**. We summarize symptoms prevalence at the point of presentation to various care settings in **Table S19**, and present attributes of cases who were tested in outpatient settings with and without symptoms in **Table S20**. Finally, in conjunction with our time-to-event analyses, we estimated aRRs for each clinical outcome using log-binomial regression models defining, as outcomes, any hospital admission or symptomatic hospital admission within 30 days of cases’ testing date, and any ICU admission, mechanical ventilation, or mortality within 60 days of cases’ testing date. Such analyses controlled for all covariates included in Cox proportional hazards models used in the primary analyses, and defined intercepts for testing date. Results comparing aHR and aRR estimates are presented in **Table S11**.

We conducted multiple (*m*=5) imputation of missing covariate values and pooled results obtained with each imputed dataset via Rubin’s rules^57^ for our primary analyses (**Table S21**). To verify our analysis results were not sensitive to the results of imputation, we compared aHR estimates for the association of Omicron vs. Delta variant infection with risk of severe outcomes from the primary analysis to results from analyses subset to cases with complete information on all measured characteristics (*N*=221,325, or 67.3% of the sample), and to results from analyses subset to cases with ≥1 year of enrollment in KPSC health plans before their diagnosis date (*N*=283,453, 86.1% of the sample), among whom fewer observations were missing (**Table S9**). To further demonstrate that missing data did not substantially affect analysis results, we also present estimates of the association of each imputed variable with the outcome of symptomatic hospital admission across the same three analysis strategies in **Table S22**, again identifying similar estimates of association in the primary analysis, in the complete-case analysis, and in the analysis subset to cases with ≥1 year of enrollment in KPSC health plans.

### Bias analysis addressing unrecorded prior infection

It has been proposed elsewhere that differential observed severity between Omicron and Delta infections may reflect that Omicron infections occur more commonly among individuals with (often unobserved) prior infection, who thus are protected by that prior infection against severe outcomes.^7^ We simulated analysis results under scenarios of differential prevalence of unobserved prior infection across case strata to determine whether our findings of reduced severity among cases with Omicron variant infection could be explained by this circumstance. We defined strata based on the joint distribution of infecting variant *i* (in recognition of reduced protection against Omicron variant infection conferred by naturally-acquired immunity from prior variants^44,45^), outcome of hospital admission or symptomatic hospital admission *j* (considering that prior infection would be expected to reduce risk of these outcomes,^17,58^ even if such protection differed by variant), and receipt of any COVID-19 vaccine doses *k* (assuming that reduced severity of infections acquired after vaccination could lead to reduced likelihood of testing and detection^59,60^). Here, defining strata based on Omicron variant infection and hospital admission status allowed us to assess how unobserved prior infections could directly impact the primary association of interest to this study. Allowing for an enhanced likelihood that prior infections among vaccinated cases went unobserved was of interest due to the higher prevalence of prior vaccination among cases with Omicron variant infection, and the possibility that the likelihood of detection of prior infection in a vaccinated individual could be reduced due to the lower severity of post-vaccination infections and relaxed requirements for SARS-CoV-2 testing as a condition for entry into workplaces and indoor public spaces among vaccinated individuals in California in 2021 (per the observed data, prevalence of documented prior infection was 0.80% and 0.38% among cases who received 0 COVID-19 vaccine doses and ≥1 COVID-19 vaccine dose, respectively). Thus, unobserved infections among vaccinated cases constituted an additional mechanism by which naturally-acquired immunity, if present, could be differentially unaccounted for in association with cases’ infecting variant.^7^

Defining *ρ_ijk_* as the observed prevalence of prior infection within any stratum, and *θ* as a multiplier conveying the proportion of infections that would be expected to go unobserved in the stratum of unvaccinated cases with Delta variant infection admitted to hospital, the probability of unobserved prior infection within the *i*, *j*, *k*^th^ stratum was *ρ_ijk_* (*θϕ_i_σ_j_ω_k_* − 1) for values of *θ* = (1, 2, 3, 4, 5), *ϕ_i_* = (1, 2, 3), *σ_j_* = (1, 2, 3), and *ω_k_* = (1, 2, 3). Within each imputed dataset, we assigned prior infection to additional cases who did not have known prior infection at random according to the probabilities *ρ_ijk_* (*θϕ_i_σ_j_ω_k_* − 1), given their observed outcome and characteristics, and repeated the primary analysis approach using stratified Cox proportional hazards models to estimate the aHR of hospital admission and symptomatic hospital admission outcomes associated with Omicron variant detection. We plot estimates of the resulting aHR for outcomes of any hospital admission and symptomatic hospital admission in **Figure S1** and **Figure S2**, respectively.

### Comparing risk of severe clinical outcomes during periods of Delta and Omicron variant predominance

To address concerns about possible bias in our primary analysis that was limited to cases tested using the ThermoFisher TaqPath COVID-19 Combo Kit, we further sought to verify whether the reduced risk of severe clinical outcomes among cases with Omicron variant infection in the primary analysis was reflected by changes severe outcomes from Delta-predominant to Omicron-predominant periods. Among all cases ascertained in outpatient settings (without restriction to cases tested using ThermoFisher TaqPath COVID-19 Combo Kit assays) over the period of 1 November, 2021 to 17 January, 2022, we fit aHRs relating the risk of severe clinical endpoints to cases’ testing date via Cox proportional hazards models. As the goal of these analyses was to relate changes in risk of severe clinical outcomes to timing of the emergence of the Omicron variant in the study population, testing dates were defined as covariates rather than as model strata, as detailed below.

Models were defined allowing for up to two changepoints in the slope of associations between testing date and risk of clinical endpoints, with changepoints defined at all dates in the study period between 15 November, 2021 and 10 January, 2022. Model formulations with zero, one, and two changepoints specified conditional hazards of each outcome given each case’s observed covariates and testing date, *λ*(*t|X_i_, τ_i_*), according to

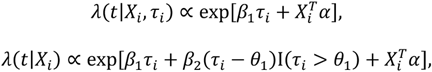

and

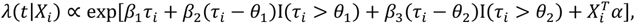

respectively. Here, *τ_i_* defines the testing date, I(*τ_i_* > *θ_k_*) serves an indicator that the testing date occurred after a changepoint in the slope at time *θ*, and 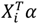is the product of all other covariates and their respective regression coefficients. We fit models defining change points at each day (or combination of days) through the time series, and used the Bayesian information criterion to define model weights:^28,61^

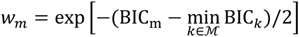

for a given model *m* from the state space of all candidate models, ℳ. Posterior model weights divided *w_m_* by the number of models fitted with the same number of change points, thereby assigning equal prior probability to models with 0, 1, or 2 changepoints. We defined testing date-specific hazards, and date-specific changepoint probabilities, by sampling models according to their posterior weights. As the sporadic occurrence of mechanical ventilation during the early study period hindered estimation of slopes in risk of this outcome, analyses addressed endpoints of hospital admission, symptomatic hospital admission, ICU admission, and death only.

### Hospital duration of stay analysis

For admitted cases diagnosed in outpatient settings with Delta or Omicron variant infection (from 15 December, 2021 to 17 January, 2022) and BA.2 or BA.1* Omicron subvariant infection (from 3 February to 17 March, 2022), we compared times from cases’ admission date to each of three possible outcomes: discharge home (without skilled care), discharge to any skilled care setting (comprising skilled nursing facilities, residential care facilities, rehabilitation facilities, other acute inpatient hospitals, or home with skilled care providers) or to home against medical advice, and in-hospital death or discharge to hospice. To compare overall rates of exit from the hospital among by infecting lineage, we additionally fit Cox proportional hazards models estimating the aHR of hospital exit (with any final disposition) associated with SGTF status, defining strata on admission date to adjust for any changes in clinical practice over time and controlling for all covariates included in the primary analysis.

### Software

We conducted all analyses using R (version 4.0.3; R Foundation for Statistical Computing, Vienna, Austria). We used the survival ^62^ package for time-to-event analyses, and the Amelia II package for multiple imputation. Analysis code is available from github.com/joelewnard/omicronSeverity. Individuals wishing to access individual-level patient data should contact the Kaiser Permanente Southern California Institutional Review Board at to enter into a data access agreement.

## Acknowledgments

This work was funded by the US Centers for Disease Control and Prevention; authors ML, RJK, and MMP are employees of this agency. The findings and conclusions in this report are those of the authors and do not necessarily represent the official position of the CDC. JAL was supported by grant R01-AI14812701A1 from the National Institute for Allergy and Infectious Diseases (US National Institutes of Health) which had no role in design or conduct of the study, or decision to submit for publication.

## Competing interests

JAL has received research grants and consulting honoraria unrelated to this study from Pfizer. SYT has received research grants unrelated to this study from Pfizer. ML has received research grants unrelated to this study from Pfizer, and has provided unpaid scientific advisory services to Janssen, Astra-Zeneca, One Day Sooner, and Covaxx (United Biomedical).

---

**Table S1:**
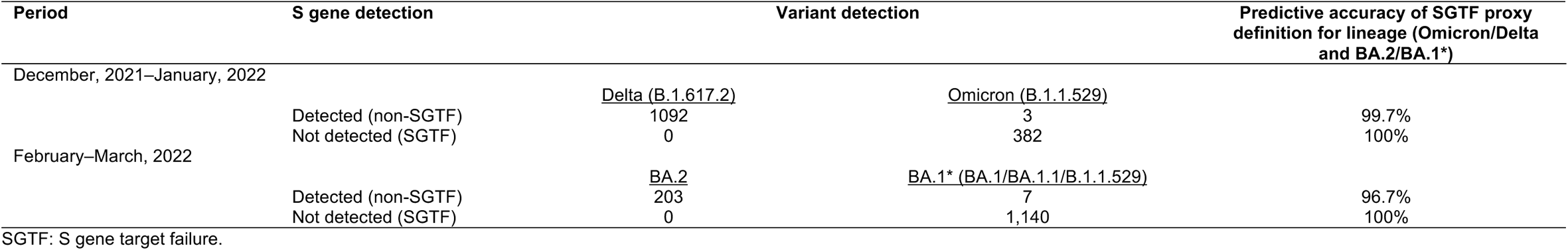
Concordance of SGTF infection status with Omicron or Delta variant detection.

**Table S2:**
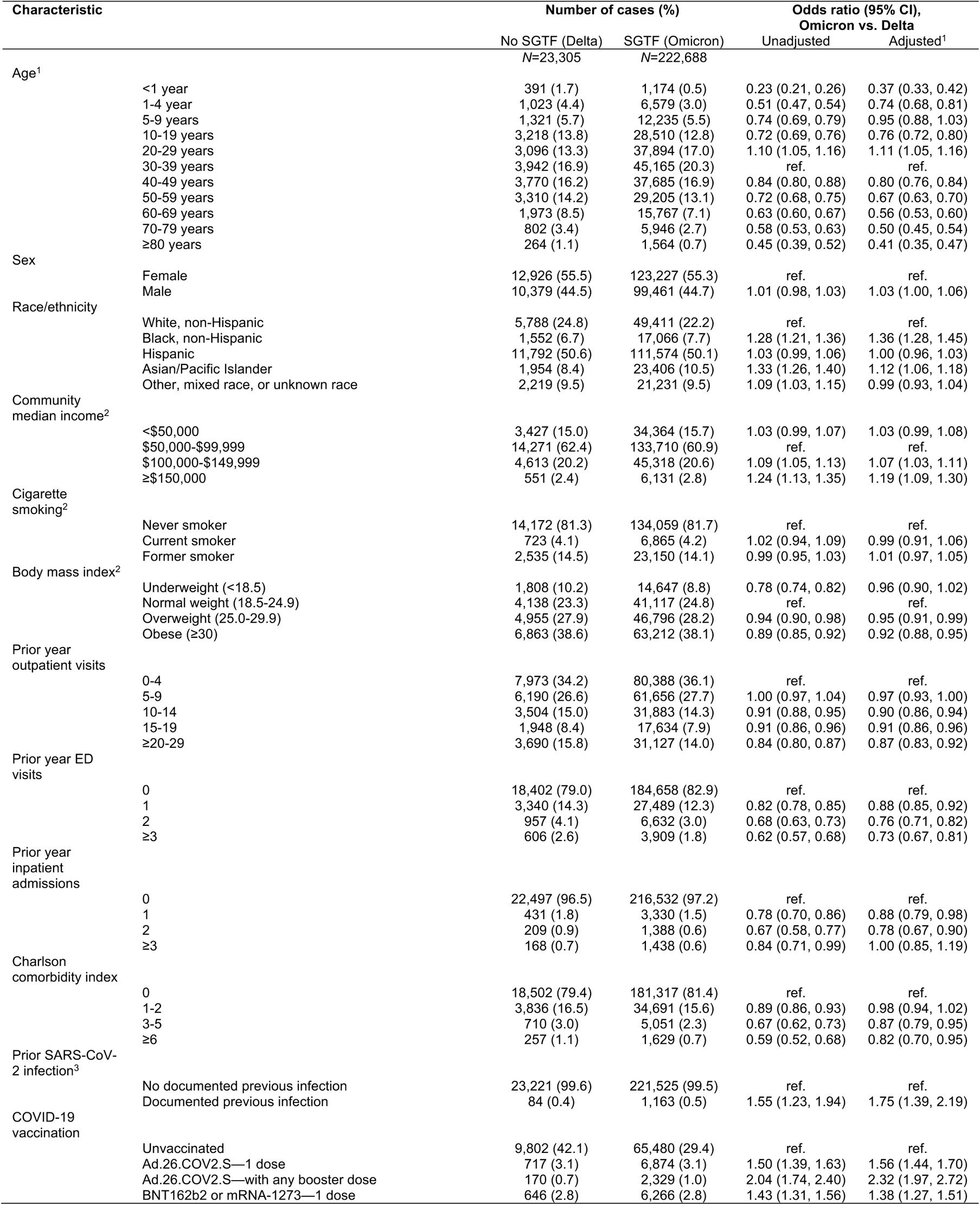
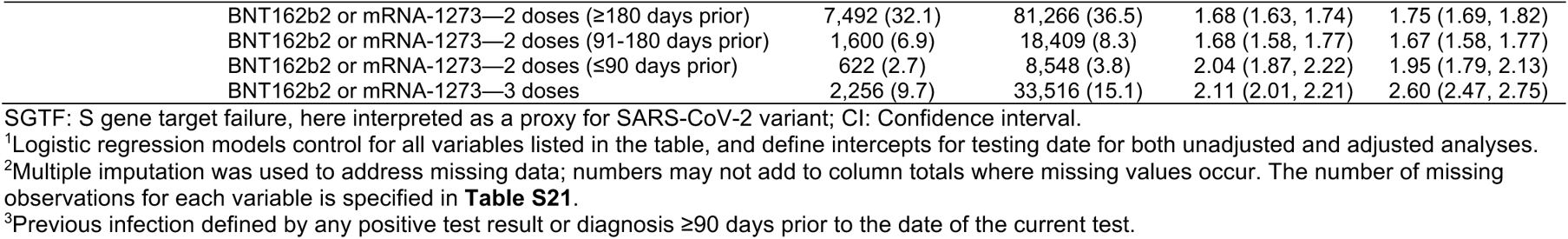
Demographic and clinical characteristics of cases tested in outpatient settings with and without SGTF, 15 December, 2021 to 17 January, 2022.

**Table S3:** Comorbid conditions among cases tested in outpatient settings with and without SGTF, 15 December, 2021 to 17 January, 2022.

| Condition | Outpatient tested cases, *n* (%) |  |  |  | Symptomatic hospitalized cases, *n* (%) |  |  |  |
| --- | --- | --- | --- | --- | --- | --- | --- | --- |
|  | No SGTF | SGTF | Odds ratio (95% CI), Omicron vs. Delta |  | No SGTF (Delta) | SGTF (Omicron) | Odds ratio (95% CI), Omicron vs. Delta |  |
|  | (Delta) | (Omicron) | Unadjusted | Adjusted<sup>1</sup> |  |  | Unadjusted | Adjusted<sup>1</sup> |
|  | *N*=23,305 | *N*=222,688 |  |  | *N*=323 | *N*=1,239 |  |  |
| Myocardial infarction | 167 (0.7) | 1,054 (0.5) | 0.60 (0.51, 0.70) | 0.87 (0.73, 1.05) | 18 (5.6) | 33 (2.7) | 0.44 (0.23, 0.84) | 0.69 (0.31, 1.53) |
| Congestive heart failure | 244 (1.0) | 1,499 (0.7) | 0.59 (0.52, 0.68) | 0.94 (0.80, 1.10) | 24 (7.4) | 74 (6.0) | 0.81 (0.48, 1.36) | 1.17 (0.60, 2.30) |
| Peripheral vascular disease | 826 (3.5) | 5,742 (2.6) | 0.67 (0.62, 0.72) | 0.97 (0.88, 1.07) | 55 (17.0) | 156 (12.6) | 0.65 (0.45, 0.94) | 1.05 (0.61, 1.80) |
| Cerebrovascular disease | 178 (0.8) | 1,273 (0.6) | 0.70 (0.59, 0.82) | 1.06 (0.89, 1.26) | 14 (4.3) | 31 (2.5) | 0.63 (0.31, 1.29) | 1.02 (0.45, 2.35) |
| Diabetes—controlled | 1,303 (5.6) | 12,004 (5.4) | 0.90 (0.85, 0.96) | 0.99 (0.92, 1.05) | 32 (9.9) | 146 (11.8) | 1.20 (0.77, 1.86) | 1.16 (0.74, 1.83) |
| Diabetes—uncontrolled | 739 (3.2) | 5,432 (2.4) | 0.71 (0.65, 0.76) | 0.92 (0.83, 1.01) | 41 (12.7) | 139 (11.2) | 0.85 (0.56, 1.27) | 0.88 (0.52, 1.47) |
| Liver disease—mild | 566 (2.4) | 4,848 (2.2) | 0.86 (0.79, 0.94) | 0.98 (0.90, 1.08) | 11 (3.4) | 67 (5.4) | 1.71 (0.85, 3.44) | 1.66 (0.84, 3.26) |
| Liver disease—moderate to severe | 32 (0.1) | 148 (0.1) | 0.46 (0.31, 0.68) | 0.72 (0.48, 1.07) | 1 (0.3) | 9 (0.7) | 1.55 (0.19, 12.8) | 1.85 (0.20, 17.1) |
| Renal disease | 515 (2.2) | 3,539 (1.6) | 0.67 (0.61, 0.73) | 0.93 (0.83, 1.05) | 33 (10.2) | 128 (10.3) | 1.06 (0.68, 1.65) | 1.41 (0.77, 2.60) |
| Chronic obstructive pulmonary disease | 1,864 (8.0) | 16,071 (7.2) | 0.88 (0.84, 0.93) | 0.98 (0.87, 1.10) | 58 (18.0) | 162 (13.1) | 0.62 (0.43, 0.88) | 0.43 (0.22, 0.86) |
| Asthma | 1,741 (7.5) | 15,219 (6.8) | 0.90 (0.85, 0.95) | 0.99 (0.88, 1.11) | 41 (12.7) | 135 (10.9) | 0.78 (0.52, 1.16) | 1.58 (0.75, 3.31) |
| Apnea | 801 (3.4) | 6,743 (3.0) | 0.86 (0.80, 0.92) | 0.99 (0.92, 1.08) | 25 (7.7) | 96 (7.7) | 0.96 (0.59, 1.58) | 1.26 (0.73, 2.16) |
| Hyperlipidemia | 3,522 (15.1) | 30,348 (13.6) | 0.85 (0.82, 0.89) | 1.02 (0.97, 1.07) | 109 (33.7) | 342 (27.6) | 0.68 (0.51, 0.91) | 0.85 (0.59, 1.22) |
| Hypothyroid | 741 (3.2) | 5,904 (2.7) | 0.81 (0.75, 0.88) | 0.95 (0.88, 1.04) | 19 (5.9) | 84 (6.8) | 1.09 (0.62, 1.89) | 1.07 (0.61, 1.88) |
| Depression | 1,868 (8.0) | 17,715 (8.0) | 1.01 (0.96, 1.06) | 1.03 (0.97, 1.09) | 37 (11.5) | 155 (12.5) | 1.25 (0.82, 1.89) | 1.21 (0.77, 1.89) |
| Dementia | 52 (0.2) | 313 (0.1) | 0.55 (0.41, 0.74) | 0.93 (0.68, 1.27) | 4 (1.2) | 14 (1.1) | 0.80 (0.24, 2.67) | 1.03 (0.26, 4.10) |
| Rheumatic disease | 203 (0.9) | 1,568 (0.7) | 0.78 (0.68, 0.91) | 0.90 (0.77, 1.05) | 3 (0.9) | 33 (2.7) | 4.01 (1.04, 15.5) | 2.67 (0.84, 8.53) |
| Peptic ulcer disease | 43 (0.2) | 216 (0.1) | 0.51 (0.36, 0.71) | 0.69 (0.49, 0.97) | 3 (0.9) | 11 (0.9) | 1.17 (0.29, 4.74) | 0.91 (0.18, 4.70) |
| Paraplegia and hemiplegia | 37 (0.2) | 272 (0.1) | 0.72 (0.51, 1.03) | 0.98 (0.68, 1.41) | 3 (0.9) | 8 (0.6) | 0.70 (0.16, 3.14) | 0.43 (0.10, 1.85) |
| Organ transplantation | 44 (0.2) | 252 (0.1) | 0.55 (0.40, 0.77) | 0.61 (0.43, 0.87) | 2 (0.6) | 28 (2.3) | 2.71 (0.63, 11.7) | 0.72 (0.18, 2.85) |
| HIV | 13 (0.1) | 133 (0.1) | 0.99 (0.56, 1.75) | 0.96 (0.54, 1.72) | 0 (0.0) | 1 (0.1) | — | — |
| Cancer—Metastatic carcinoma | 74 (0.3) | 592 (0.3) | 0.81 (0.63, 1.04) | 1.05 (0.81, 1.35) | 4 (1.2) | 14 (1.1) | 0.82 (0.25, 2.70) | 1.18 (0.35, 3.93) |
| Cancer—other | 289 (1.2) | 2,234 (1.0) | 0.77 (0.68, 0.88) | 0.96 (0.84, 1.09) | 13 (4.0) | 59 (4.8) | 1.09 (0.57, 2.07) | 1.17 (0.61, 2.23) |
| Other immunocompromising or immune suppressing condition | 541 (2.3) | 4,778 (2.1) | 0.91 (0.84, 1.00) | 1.04 (0.95, 1.15) | 17 (5.3) | 91 (7.3) | 1.55 (0.87, 2.75) | 1.45 (0.77, 2.72) |

SGTF: S gene target failure, here interpreted as a proxy for SARS-CoV-2 variant; CI: Confidence interval.
<sup>1</sup>Logistic regression models control for all variables listed in **Table S2**, and define intercepts for testing date for both unadjusted and adjusted analyses.

**Table S4:**
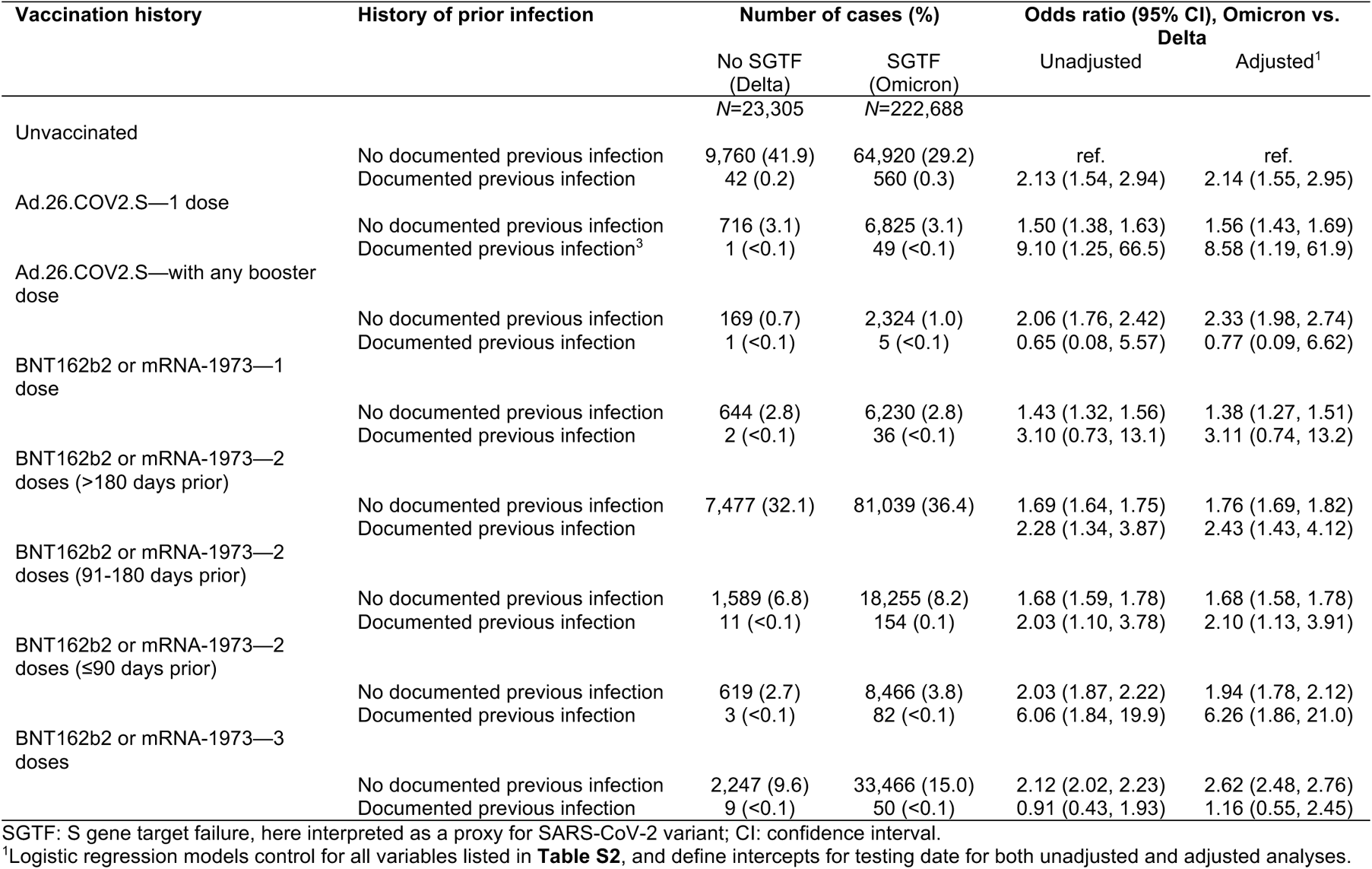
Combined history of documented SARS-CoV-2 infection and COVID-19 vaccination among cases diagnosed in outpatient settings with and without SGTF, 15 December, 2021 to 17 January, 2022.

**Table S5:**
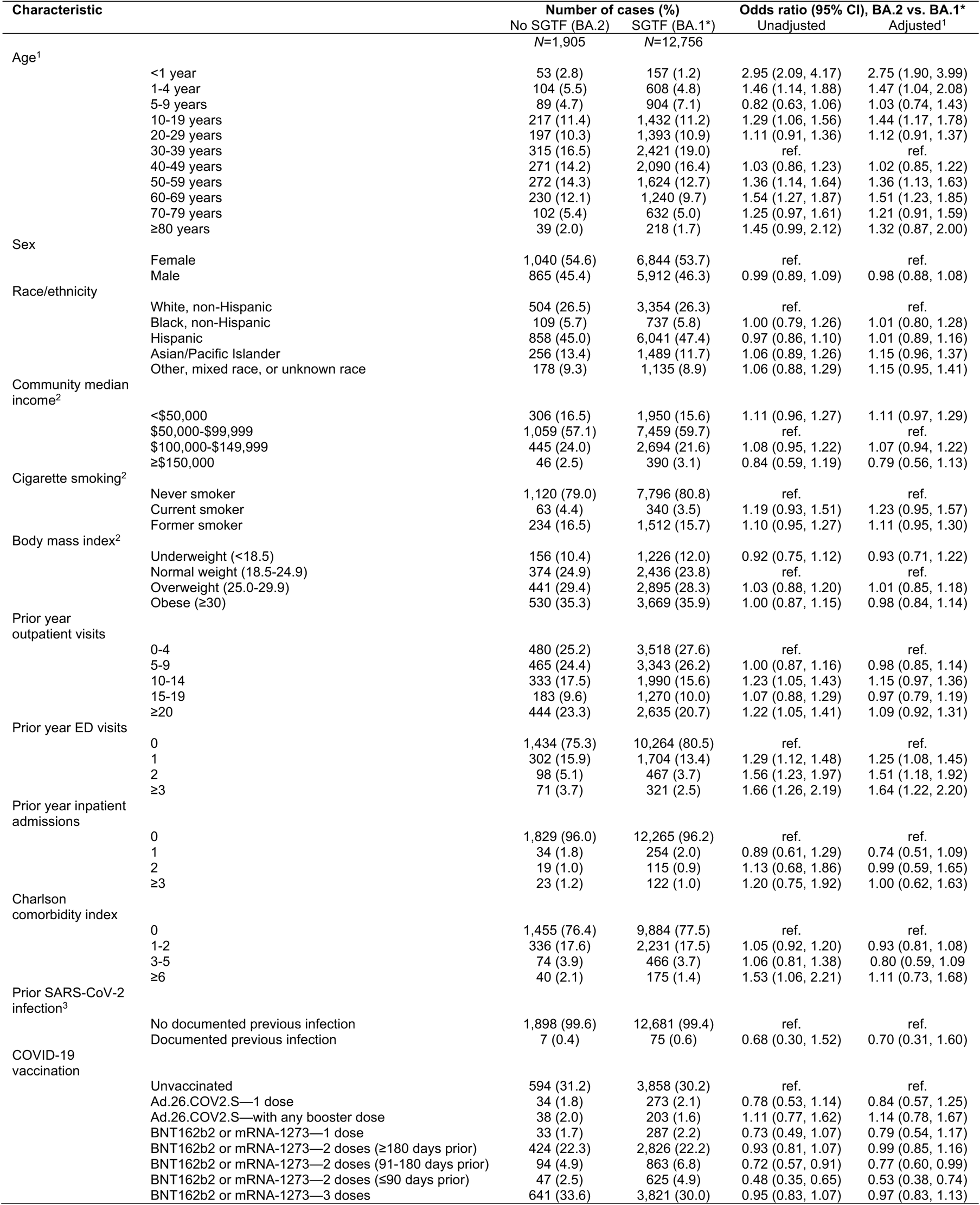
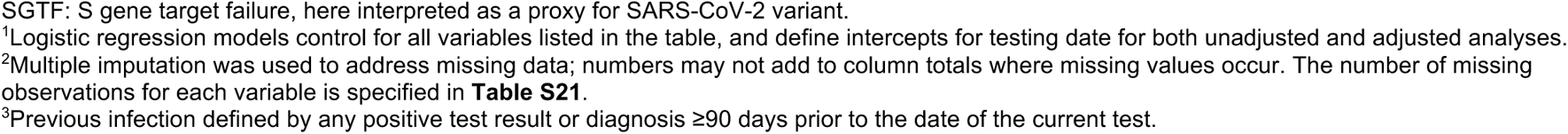
Demographic and clinical characteristics of cases tested in outpatient settings with and without SGTF, 3 February to 17 March, 2022.

| Characteristic |  | Number of cases (%) |  | Odds ratio (95% CI), BA.2 vs. BA.1* |  |
| --- | --- | --- | --- | --- | --- |
|  |  | No SGTF (BA.2)<br>N=1,905 | SGTF (BA.1*)<br>N=12,756 | Unadjusted | Adjusted<sup>1</sup> |
| Age<sup>1</sup> | <1 year | 53 (2.8) | 157 (1.2) | 2.95 (2.09, 4.17) | 2.75 (1.90, 3.99) |
|  | 1-4 year | 104 (5.5) | 608 (4.8) | 1.46 (1.14, 1.88) | 1.47 (1.04, 2.08) |
|  | 5-9 years | 89 (4.7) | 904 (7.1) | 0.82 (0.63, 1.06) | 1.03 (0.74, 1.43) |
|  | 10-19 years | 217 (11.4) | 1,432 (11.2) | 1.29 (1.06, 1.56) | 1.44 (1.17, 1.78) |
|  | 20-29 years | 197 (10.3) | 1,393 (10.9) | 1.11 (0.91, 1.36) | 1.12 (0.91, 1.37) |
|  | 30-39 years | 315 (16.5) | 2,421 (19.0) | ref. | ref. |
|  | 40-49 years | 271 (14.2) | 2,090 (16.4) | 1.03 (0.86, 1.23) | 1.02 (0.85, 1.22) |
|  | 50-59 years | 272 (14.3) | 1,624 (12.7) | 1.36 (1.14, 1.64) | 1.36 (1.13, 1.63) |
|  | 60-69 years | 230 (12.1) | 1,240 (9.7) | 1.54 (1.27, 1.87) | 1.51 (1.23, 1.85) |
|  | 70-79 years | 102 (5.4) | 632 (5.0) | 1.25 (0.97, 1.61) | 1.21 (0.91, 1.59) |
|  | ≥80 years | 39 (2.0) | 218 (1.7) | 1.45 (0.99, 2.12) | 1.32 (0.87, 2.00) |
| Sex | Female | 1,040 (54.6) | 6,844 (53.7) | ref. | ref. |
|  | Male | 865 (45.4) | 5,912 (46.3) | 0.99 (0.89, 1.09) | 0.98 (0.88, 1.08) |
| Race/ethnicity | White, non-Hispanic | 504 (26.5) | 3,354 (26.3) | ref. | ref. |
|  | Black, non-Hispanic | 109 (5.7) | 737 (5.8) | 1.00 (0.79, 1.26) | 1.01 (0.80, 1.28) |
|  | Hispanic | 858 (45.0) | 6,041 (47.4) | 0.97 (0.86, 1.10) | 1.01 (0.89, 1.16) |
|  | Asian/Pacific Islander | 256 (13.4) | 1,489 (11.7) | 1.06 (0.89, 1.26) | 1.15 (0.96, 1.37) |
|  | Other, mixed race, or unknown race | 178 (9.3) | 1,135 (8.9) | 1.06 (0.88, 1.29) | 1.15 (0.95, 1.41) |
| Community median income<sup>2</sup> | <\$50,000 | 306 (16.5) | 1,950 (15.6) | 1.11 (0.96, 1.27) | 1.11 (0.97, 1.29) |
| | \$50,000-\$99,999 | 1,059 (57.1) | 7,459 (59.7) | ref. | ref. |
| | \$100,000-\$149,999 | 445 (24.0) | 2,694 (21.6) | 1.08 (0.95, 1.22) | 1.07 (0.94, 1.22) |
| | ≥\$150,000 | 46 (2.5) | 390 (3.1) | 0.84 (0.59, 1.19) | 0.79 (0.56, 1.13) |
| Cigarette smoking<sup>2</sup> | Never smoker | 1,120 (79.0) | 7,796 (80.8) | ref. | ref. |
|  | Current smoker | 63 (4.4) | 340 (3.5) | 1.19 (0.93, 1.51) | 1.23 (0.95, 1.57) |
|  | Former smoker | 234 (16.5) | 1,512 (15.7) | 1.10 (0.95, 1.27) | 1.11 (0.95, 1.30) |
| Body mass index<sup>2</sup> | Underweight (<18.5) | 156 (10.4) | 1,226 (12.0) | 0.92 (0.75, 1.12) | 0.93 (0.71, 1.22) |
|  | Normal weight (18.5-24.9) | 374 (24.9) | 2,436 (23.8) | ref. | ref. |
|  | Overweight (25.0-29.9) | 441 (29.4) | 2,895 (28.3) | 1.03 (0.88, 1.20) | 1.01 (0.85, 1.18) |
|  | Obese (≥30) | 530 (35.3) | 3,669 (35.9) | 1.00 (0.87, 1.15) | 0.98 (0.84, 1.14) |
| Prior year outpatient visits | 0-4 | 480 (25.2) | 3,518 (27.6) | ref. | ref. |
|  | 5-9 | 465 (24.4) | 3,343 (26.2) | 1.00 (0.87, 1.16) | 0.98 (0.85, 1.14) |
|  | 10-14 | 333 (17.5) | 1,990 (15.6) | 1.23 (1.05, 1.43) | 1.15 (0.97, 1.36) |
|  | 15-19 | 183 (9.6) | 1,270 (10.0) | 1.07 (0.88, 1.29) | 0.97 (0.79, 1.19) |
|  | ≥20 | 444 (23.3) | 2,635 (20.7) | 1.22 (1.05, 1.41) | 1.09 (0.92, 1.31) |
| Prior year ED visits | 0 | 1,434 (75.3) | 10,264 (80.5) | ref. | ref. |
|  | 1 | 302 (15.9) | 1,704 (13.4) | 1.29 (1.12, 1.48) | 1.25 (1.08, 1.45) |
|  | 2 | 98 (5.1) | 467 (3.7) | 1.56 (1.23, 1.97) | 1.51 (1.18, 1.92) |
|  | ≥3 | 71 (3.7) | 321 (2.5) | 1.66 (1.26, 2.19) | 1.64 (1.22, 2.20) |
| Prior year inpatient admissions | 0 | 1,829 (96.0) | 12,265 (96.2) | ref. | ref. |
|  | 1 | 34 (1.8) | 254 (2.0) | 0.89 (0.61, 1.29) | 0.74 (0.51, 1.09) |
|  | 2 | 19 (1.0) | 115 (0.9) | 1.13 (0.68, 1.86) | 0.99 (0.59, 1.65) |
|  | ≥3 | 23 (1.2) | 122 (1.0) | 1.20 (0.75, 1.92) | 1.00 (0.62, 1.63) |
| Charlson comorbidity index | 0 | 1,455 (76.4) | 9,884 (77.5) | ref. | ref. |
|  | 1-2 | 336 (17.6) | 2,231 (17.5) | 1.05 (0.92, 1.20) | 0.93 (0.81, 1.08) |
|  | 3-5 | 74 (3.9) | 466 (3.7) | 1.06 (0.81, 1.38) | 0.80 (0.59, 1.09) |
|  | ≥6 | 40 (2.1) | 175 (1.4) | 1.53 (1.06, 2.21) | 1.11 (0.73, 1.68) |
| Prior SARS-CoV-2 infection<sup>3</sup> | No documented previous infection | 1,898 (99.6) | 12,681 (99.4) | ref. | ref. |
|  | Documented previous infection | 7 (0.4) | 75 (0.6) | 0.68 (0.30, 1.52) | 0.70 (0.31, 1.60) |
| COVID-19 vaccination | Unvaccinated | 594 (31.2) | 3,858 (30.2) | ref. | ref. |
|  | Ad.26.COV2.S—1 dose | 34 (1.8) | 273 (2.1) | 0.78 (0.53, 1.14) | 0.84 (0.57, 1.25) |
|  | Ad.26.COV2.S—with any booster dose | 38 (2.0) | 203 (1.6) | 1.11 (0.77, 1.62) | 1.14 (0.78, 1.67) |
|  | BNT162b2 or mRNA-1273—1 dose | 33 (1.7) | 287 (2.2) | 0.73 (0.49, 1.07) | 0.79 (0.54, 1.17) |
|  | BNT162b2 or mRNA-1273—2 doses (≥180 days prior) | 424 (22.3) | 2,826 (22.2) | 0.93 (0.81, 1.07) | 0.99 (0.85, 1.16) |
|  | BNT162b2 or mRNA-1273—2 doses (91-180 days prior) | 94 (4.9) | 863 (6.8) | 0.72 (0.57, 0.91) | 0.77 (0.60, 0.99) |
|  | BNT162b2 or mRNA-1273—2 doses (≤90 days prior) | 47 (2.5) | 625 (4.9) | 0.48 (0.35, 0.65) | 0.53 (0.38, 0.74) |
|  | BNT162b2 or mRNA-1273—3 doses | 641 (33.6) | 3,821 (30.0) | 0.95 (0.83, 1.07) | 0.97 (0.83, 1.13) |

SGTF: S gene target failure, here interpreted as a proxy for SARS-CoV-2 variant.
<sup>1</sup>Logistic regression models control for all variables listed in the table, and define intercepts for testing date for both unadjusted and adjusted analyses.
<sup>2</sup>Multiple imputation was used to address missing data; numbers may not add to column totals where missing values occur. The number of missing observations for each variable is specified in **Table S21**.
<sup>3</sup>Previous infection defined by any positive test result or diagnosis $\geq 90$ days prior to the date of the current test.

**Table S6:**
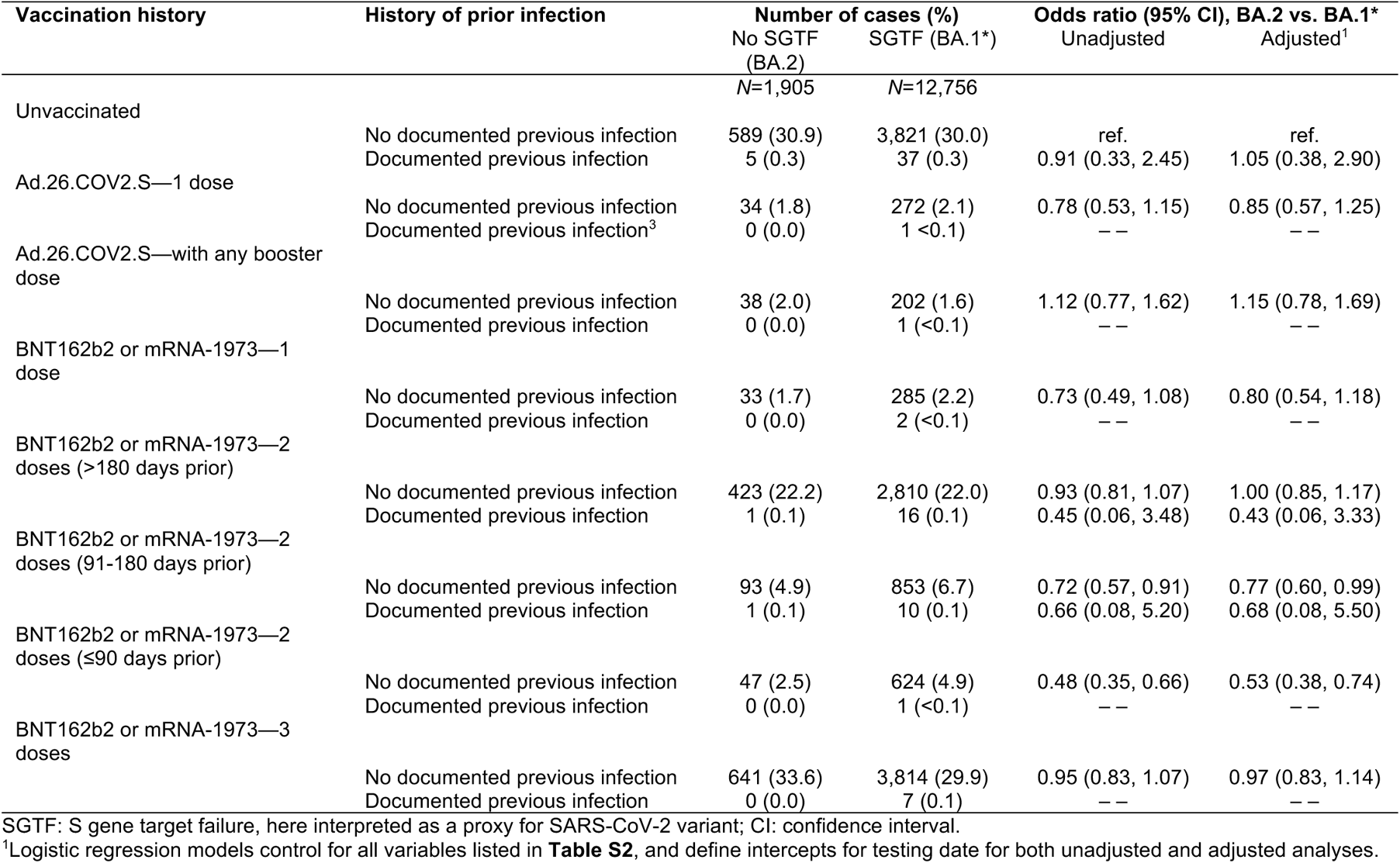
Combined history of documented SARS-CoV-2 infection and COVID-19 vaccination among cases diagnosed in outpatient settings with and without SGTF, 3 February to 17 March, 2022.

| Vaccination history | History of prior infection | Number of cases (%) |  | Odds ratio (95% CI), BA.2 vs. BA.1* |  |
| --- | --- | --- | --- | --- | --- |
|  |  | No SGTF (BA.2) | SGTF (BA.1*) | Unadjusted | Adjusted<sup>1</sup> |
|  |  | N=1,905 | N=12,756 |  |  |
| Unvaccinated | No documented previous infection | 589 (30.9) | 3,821 (30.0) | ref. | ref. |
|  | Documented previous infection | 5 (0.3) | 37 (0.3) | 0.91 (0.33, 2.45) | 1.05 (0.38, 2.90) |
| Ad.26.COV2.S—1 dose | No documented previous infection | 34 (1.8) | 272 (2.1) | 0.78 (0.53, 1.15) | 0.85 (0.57, 1.25) |
|  | Documented previous infection<sup>3</sup> | 0 (0.0) | 1 (<0.1) | -- | -- |
| Ad.26.COV2.S—with any booster dose | No documented previous infection | 38 (2.0) | 202 (1.6) | 1.12 (0.77, 1.62) | 1.15 (0.78, 1.69) |
|  | Documented previous infection | 0 (0.0) | 1 (<0.1) | -- | -- |
| BNT162b2 or mRNA-1973—1 dose | No documented previous infection | 33 (1.7) | 285 (2.2) | 0.73 (0.49, 1.08) | 0.80 (0.54, 1.18) |
|  | Documented previous infection | 0 (0.0) | 2 (<0.1) | -- | -- |
| BNT162b2 or mRNA-1973—2 doses (>180 days prior) | No documented previous infection | 423 (22.2) | 2,810 (22.0) | 0.93 (0.81, 1.07) | 1.00 (0.85, 1.17) |
|  | Documented previous infection | 1 (0.1) | 16 (0.1) | 0.45 (0.06, 3.48) | 0.43 (0.06, 3.33) |
| BNT162b2 or mRNA-1973—2 doses (91-180 days prior) | No documented previous infection | 93 (4.9) | 853 (6.7) | 0.72 (0.57, 0.91) | 0.77 (0.60, 0.99) |
|  | Documented previous infection | 1 (0.1) | 10 (0.1) | 0.66 (0.08, 5.20) | 0.68 (0.08, 5.50) |
| BNT162b2 or mRNA-1973—2 doses (≤90 days prior) | No documented previous infection | 47 (2.5) | 624 (4.9) | 0.48 (0.35, 0.66) | 0.53 (0.38, 0.74) |
|  | Documented previous infection | 0 (0.0) | 1 (<0.1) | -- | -- |
| BNT162b2 or mRNA-1973—3 doses | No documented previous infection | 641 (33.6) | 3,814 (29.9) | 0.95 (0.83, 1.07) | 0.97 (0.83, 1.14) |
|  | Documented previous infection | 0 (0.0) | 7 (0.1) | -- | -- |

SGTF: S gene target failure, here interpreted as a proxy for SARS-CoV-2 variant; CI: confidence interval.
<sup>1</sup>Logistic regression models control for all variables listed in **Table S2**, and define intercepts for testing date for both unadjusted and adjusted analyses.

**Table S7:** Occurrence of severe clinical outcomes among cases with and without SGTF, 15 December, 2021 to 17 January, 2022.

| Outcome | Case population<sup>1</sup> | Cumulative events over observed follow-up (*n* of events per 1000 cases) |  | Event rate per 10,000 person-days observed (*n* of events/person-days at risk) |  | Hazard ratio (95% CI), Omicron vs. Delta |  |
| --- | --- | --- | --- | --- | --- | --- | --- |
|  |  | No SGTF (Delta) | SGTF (Omicron) | No SGTF (Delta) | SGTF (Omicron) | Unadjusted | Adjusted |
| Any hospital admission | Cases tested in outpatient settings | 241 (10.3) | 991 (4.5) | 3.61 (241/667,630) | 1.55 (991/6,413,416) | 0.48 (0.42, 0.56) | 0.59 (0.51, 0.69) |
|  | All cases | 369 (15.7) | 1,642 (7.3) | 5.53 (369/667,696) | 2.56 (1,642/6,413,759) | 0.50 (0.44, 0.56) | 0.61 (0.54, 0.68) |
| Symptomatic hospital admission<sup>1</sup> | Cases tested in outpatient settings | 226 (9.7) | 878 (3.9) | 3.39 (226/667,630) | 1.37 (878/6,413,416) | 0.46 (0.40, 0.54) | 0.59 (0.51, 0.68) |
|  | All cases | 323 (13.7) | 1,239 (5.5) | 4.84 (323/667,696) | 1.93 (1,239/6,413,759) | 0.44 (0.38, 0.50) | 0.55 (0.49, 0.62) |
| ICU admission | Cases tested in outpatient settings | 26 (1.1) | 51 (0.2) | 0.19 (26/1,363,726) | 0.04 (51/13,062,884) | 0.27 (0.15, 0.40) | 0.50 (0.29, 0.87) |
|  | All cases | 29 (1.2) | 57 (0.3) | 0.21 (29/1,371,050) | 0.04 (57/13,100,749) | 0.24 (0.15, 0.39) | 0.48 (0.29, 0.81) |
| Mechanical ventilation | Cases tested in outpatient settings | 17 (0.7) | 24 (0.1) | 0.12 (17/1,364,195) | 0.02 (24/13,064,223) | 0.18 (0.09, 0.35) | 0.36 (0.18, 0.72) |
|  | All cases | 19 (0.8) | 26 (0.1) | 0.14 (19/1,371,577) | 0.02 (26/13,102,322) | 0.17 (0.09, 0.33) | 0.32 (0.17, 0.62) |
| Death | Cases tested in outpatient settings | 19 (0.8) | 19 (0.1) | 0.14 (19/1,364,392) | 0.01 (19/13,064,699) | 0.12 (0.06, 0.23) | 0.21 (0.10, 0.44) |
|  | All cases | 19 (0.8) | 19 (0.1) | 0.14 (19/1,371,886) | 0.01 (19/13,102,914) | 0.12 (0.06, 0.23) | 0.21 (0.10, 0.44) |

SGTF: S gene target failure, here interpreted as a proxy for SARS-CoV-2 variant; CI: confidence interval. Sample includes 23,305 and 222,688 cases with Delta and Omicron variant infection, respectively, diagnosed in outpatient settings.
<sup>1</sup>We define symptomatic hospital admissions as those occurring among cases with respiratory symptom onset dates on or ≤14 days before the date of admission.

**Table S8:** Occurrence of clinical outcomes among cases with and without SGTF, without symptoms at the time of their first positive test, 15 December, 2021 to 17 January, 2022.

| Outcome | Case population<sup>1</sup> | Cumulative events over observed follow-up (n of events per 1,000 cases) |  | Event rate per 10,000 person-days observed (n of events/person-days at risk) |  | Hazard ratio (95% CI), Omicron vs. Delta |  |
| --- | --- | --- | --- | --- | --- | --- | --- |
|  |  | No SGTF (Delta) | SGTF (Omicron) | No SGTF (Delta) | SGTF (Omicron) | Unadjusted | Adjusted |
| Onset of respiratory symptoms | Cases tested in outpatient settings without symptoms | 574 (13.3) | 5,720 (11.6) | 0.20 (574/280,118) | 0.18 (5,720/3,109,842) | 0.95 (0.87, 1.04) | 0.88 (0.81, 0.96) |
|  | All cases without symptoms at point of testing | 581 (13.4) | 5,787 (11.7) | 0.21 (581/281,047) | 0.19 (5,787/3,119,788) | 0.95 (0.87, 1.04) | 0.89 (0.81, 0.97) |
| Any hospital admission | Cases tested in outpatient settings without symptoms | 28 (6.5) | 179 (3.6) | 2.27 (28/123,523) | 1.26 (179/1,416,427) | 0.57 (0.38, 0.86) | 0.69 (0.46, 1.05) |
|  | All cases without symptoms at point of testing | 45 (10.4) | 356 (7.2) | 3.64 (45/123,532) | 2.51 (356/1,416,516) | 0.71 (0.52, 0.98) | 0.83 (0.60, 1.15) |
| Symptomatic hospital admission<sup>1</sup> | Cases tested in outpatient settings without symptoms | 13 (3.0) | 66 (1.3) | 1.05 (13/123,523) | 0.47 (66/1,416,427) | 0.53 (0.35, 0.80) | 0.65 (0.43, 0.98) |
|  | All cases without symptoms at point of testing | 13 (3.0) | 66 (1.3) | 1.05 (13/123,532) | 0.47 (66/1,416,516) | 0.53 (0.35, 0.80) | 0.65 (0.43, 0.98) |
| ICU admission | Cases tested in outpatient settings without symptoms | 1 (0.2) | 8 (0.2) | 0.04 (1/251,816) | 0.03 (8/2,885,008) | 0.59 (0.07, 4.70) | -- |
|  | All cases without symptoms at point of testing | 1 (0.2) | 8 (0.2) | 0.04 (1/252,819) | 0.03 (8/2,895,452) | 0.59 (0.07, 4.71) | -- |
| Mechanical ventilation | Cases tested in outpatient settings without symptoms | 0 (0.0) | 4 (0.1) | 0.00 (0/251,978) | 0.01 (4/2,885,243) | -- | -- |
|  | All cases without symptoms at point of testing | 0 (0.0) | 4 (0.1) | 0.00 (0/252,981) | 0.01 (4/2,895,686) | -- | -- |
| Death | Cases tested in outpatient settings without symptoms | 0 (0.0) | 0 (0.0) | 0.00 (0/251,978) | 0.00 (0/2,885,381) | -- | -- |
|  | All cases without symptoms at point of testing | 0 (0.0) | 0 (0.0) | 0.00 (0/252,981) | 0.00 (0/2,895,824) | -- | -- |

SGTF: S gene target failure, here interpreted as a proxy for SARS-CoV-2 variant; CI: confidence interval.
Symptoms data were collected from free-text fields of patient EHRs, including data from clinical notes and from responses to questionnaires addressing symptoms in the 14 days before testing which all patients must complete to receive a test. Note that because lengths of follow-up differ between SGTF and non-SGTF cases, clinical risk periods over which cumulative events and event rates are summarized may differ between the two case populations.
<sup>1</sup>We define symptomatic hospital admissions as those occurring among cases with respiratory symptom onset dates on or ≤14 days before the date of admission.

**Table S9:** Comparison of associations of SGTF with clinical outcomes across differing missing-data analysis strategies, 15 December, 2021 to 17 January, 2022.

| Outcome | Case population<sup>1</sup> | Primary analysis: Adjusted hazard ratio (95% CI), Omicron vs. Delta | Complete case analysis: Adjusted hazard ratio (95% CI), Omicron vs. Delta | Participants enrolled ≥1 year: Adjusted hazard ratio (95% CI), Omicron vs. Delta |
| --- | --- | --- | --- | --- |
| Any hospital admission | Cases tested in outpatient settings | 0.59 (0.51, 0.69) | 0.65 (0.55, 0.76) | 0.59 (0.51, 0.70) |
|  | All cases | 0.61 (0.54, 0.68) | 0.66 (0.58, 0.75) | 0.62 (0.55, 0.70) |
| Symptomatic hospital admission<sup>1</sup> | Cases tested in outpatient settings | 0.59 (0.51, 0.68) | 0.64 (0.54, 0.75) | 0.59 (0.50, 0.69) |
|  | All cases | 0.55 (0.49, 0.62) | 0.61 (0.53, 0.70) | 0.56 (0.49, 0.64) |
| ICU admission | Cases tested in outpatient settings | 0.50 (0.29, 0.87) | 0.53 (0.30, 0.95) | 0.51 (0.29, 0.91) |
|  | All cases | 0.48 (0.29, 0.81) | 0.50 (0.29, 0.87) | 0.52 (0.29, 0.92) |
| Mechanical ventilation | Cases tested in outpatient settings | 0.36 (0.18, 0.72) | 0.32 (0.16, 0.65) | 0.32 (0.16, 0.65) |
|  | All cases | 0.32 (0.17, 0.62) | 0.30 (0.15, 0.61) | 0.34 (0.17, 0.68) |
| Death | Cases tested in outpatient settings | 0.21 (0.10, 0.44) | 0.18 (0.08, 0.39) | 0.20 (0.09, 0.42) |
|  | All cases | 0.21 (0.10, 0.44) | 0.18 (0.08, 0.40) | 0.20 (0.09, 0.43) |

SGTF: S gene target failure, here interpreted as a proxy for SARS-CoV-2 variant; CI: confidence interval. Adjusted Cox proportional hazards models control for all variables listed in **Table S2**, with strata for testing date.
<sup>1</sup>We define symptomatic hospital admissions as those occurring among cases with respiratory symptom onset dates on or ≤14 days before the date of admission.

**Table S10:**
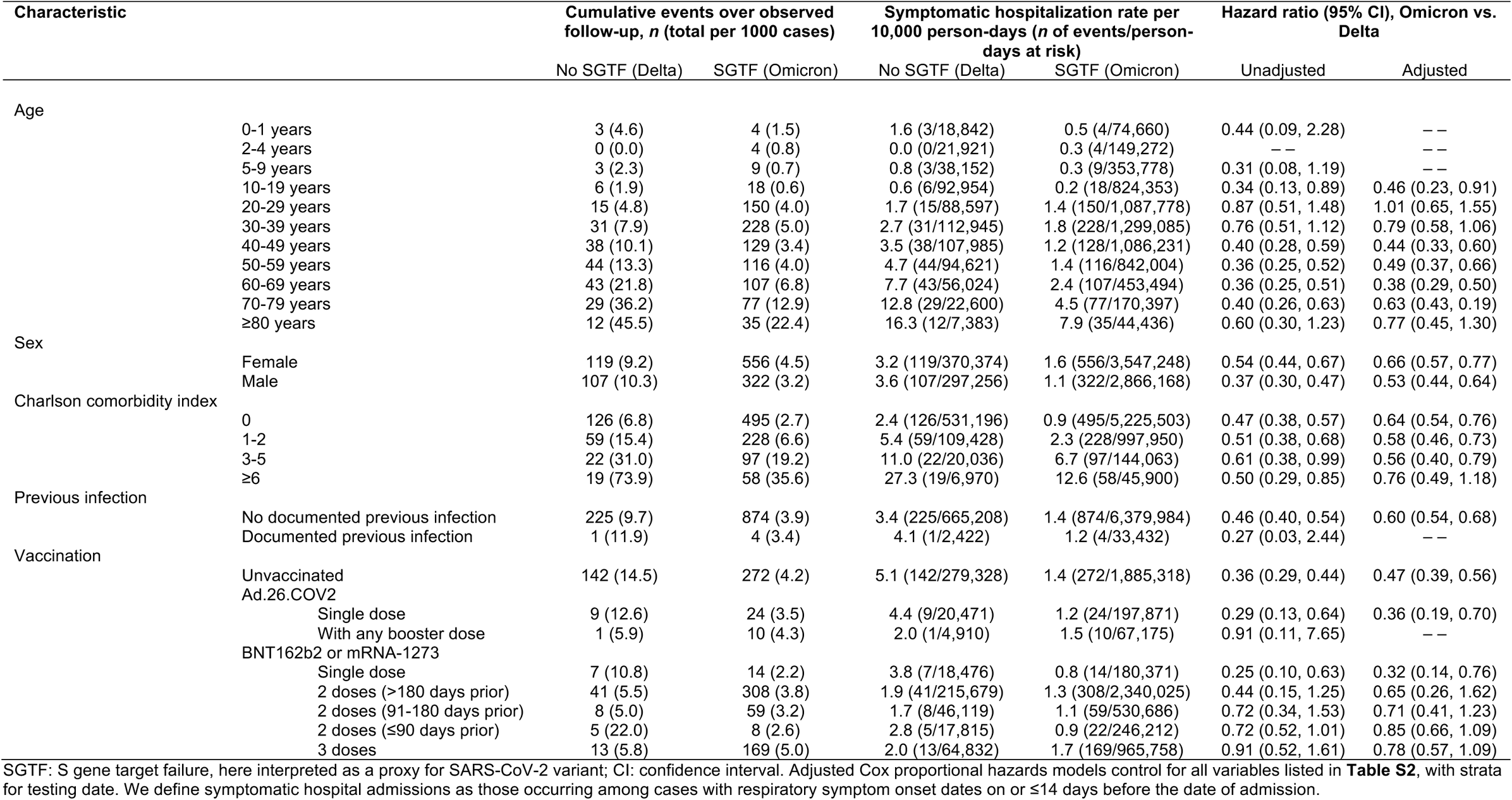
Association of SGTF infection status with symptomatic hospitalization within various patient subgroups, 15 December, 2021 to 17 January, 2022.

**Table S11:** Comparison of associations of SGTF with clinical outcomes defined on the basis of the adjusted hazard ratio or adjusted risk ratio, 15 December, 2021 to 17 January, 2022.

| Outcome | Case population<sup>1</sup> | Primary analysis: Adjusted hazard ratio (95% CI), Omicron vs. Delta | Log binomial regression analysis: Risk ratio (95% CI), Omicron vs. Delta |
| --- | --- | --- | --- |
| Any hospital admission | Cases tested in outpatient settings | 0.59 (0.51, 0.69) | 0.63 (0.55, 0.72) |
|  | All cases | 0.61 (0.54, 0.68) | 0.67 (0.60, 0.75) |
| Symptomatic hospital admission<sup>1</sup> | Cases tested in outpatient settings | 0.59 (0.51, 0.68) | 0.63 (0.55, 0.72) |
|  | All cases | 0.55 (0.49, 0.62) | 0.60 (0.54, 0.68) |
| ICU admission | Cases tested in outpatient settings | 0.50 (0.29, 0.87) | 0.54 (0.31, 0.94) |
|  | All cases | 0.48 (0.29, 0.81) | 0.51 (0.30, 0.86) |
| Mechanical ventilation | Cases tested in outpatient settings | 0.36 (0.18, 0.72) | 0.35 (0.17, 0.71) |
|  | All cases | 0.32 (0.17, 0.62) | 0.32 (0.16, 0.62) |
| Death | Cases tested in outpatient settings | 0.21 (0.10, 0.44) | 0.20 (0.10, 0.43) |
|  | All cases | 0.21 (0.10, 0.44) | 0.21 (0.10, 0.43) |

SGTF: S gene target failure, here interpreted as a proxy for SARS-CoV-2 variant; CI: confidence interval. Adjusted Cox proportional hazards and log binomial regression models control for all variables listed in **Table S2**, with strata and intercepts for testing date, respectively.
<sup>1</sup>We define symptomatic hospital admissions as those occurring among cases with respiratory symptom onset dates on or ≤14 days before the date of admission.

**Table S12:** Occurrence of severe clinical outcomes among cases tested in outpatient settings with and without SGTF, 3 February to 17 March, 2022.

| Outcome | Cumulative events over observed follow-up<br>(*n* of events per 1000 cases) |  | Event rate per 10,000 person-days observed (*n* of<br>events/person-days at risk) |  | Hazard ratio (95% CI), BA.2 vs. BA.1* |  |
| --- | --- | --- | --- | --- | --- | --- |
|  | No SGTF (BA.2) | SGTF (BA.1*) | No SGTF (BA.2) | SGTF (BA.1*) | Unadjusted | Adjusted |
| Any hospital admission | 28 (14.7) | 170 (13.3) | 7.01 (28/39,965) | 5.16 (170/329,302) | 1.38 (0.92, 2.07) | 1.26 (0.83, 1.92) |
| Symptomatic hospital admission<sup>1</sup> | 24 (12.6) | 147 (11.5) | 6.01 (24/39,965) | 4.46 (147/329,302) | 1.42 (0.92, 2.20) | 1.29 (0.82, 2.03) |
| ICU admission | 1 (0.5) | 6 (0.5) | 0.21 (1/48,374) | 0.15 (6/406,887) | 1.63 (0.20, 13.8) | -- |
| Mechanical ventilation | 1 (0.5) | 1 (0.1) | 0.21 (1/48,374) | 0.02 (1/406,982) | -- | -- |
| Death | 2 (1.0) | 16 (1.3) | 0.41 (2/48,367) | 0.39 (16/406,769) | 1.04 (0.24, 4.55) | 1.16 (0.25, 5.29) |

SGTF: S gene target failure, here interpreted as a proxy for SARS-CoV-2 variant; CI: confidence interval. Adjusted Cox proportional hazards models control for all variables listed in **Table S2**, with strata for testing date.
<sup>1</sup>We define symptomatic hospital admissions as those occurring among cases with respiratory symptom onset dates on or ≤14 days before the date of admission.

**Table S13:**
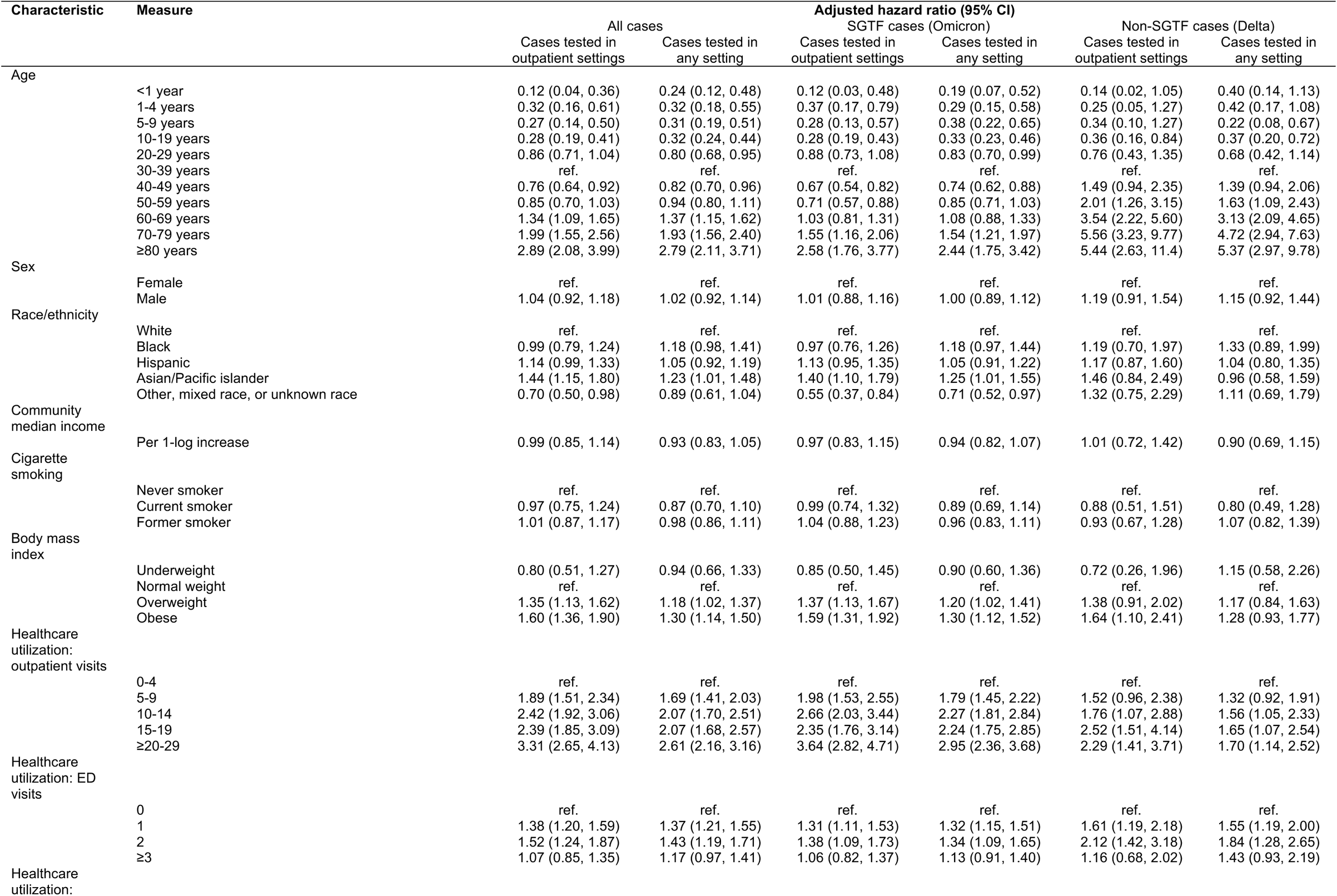
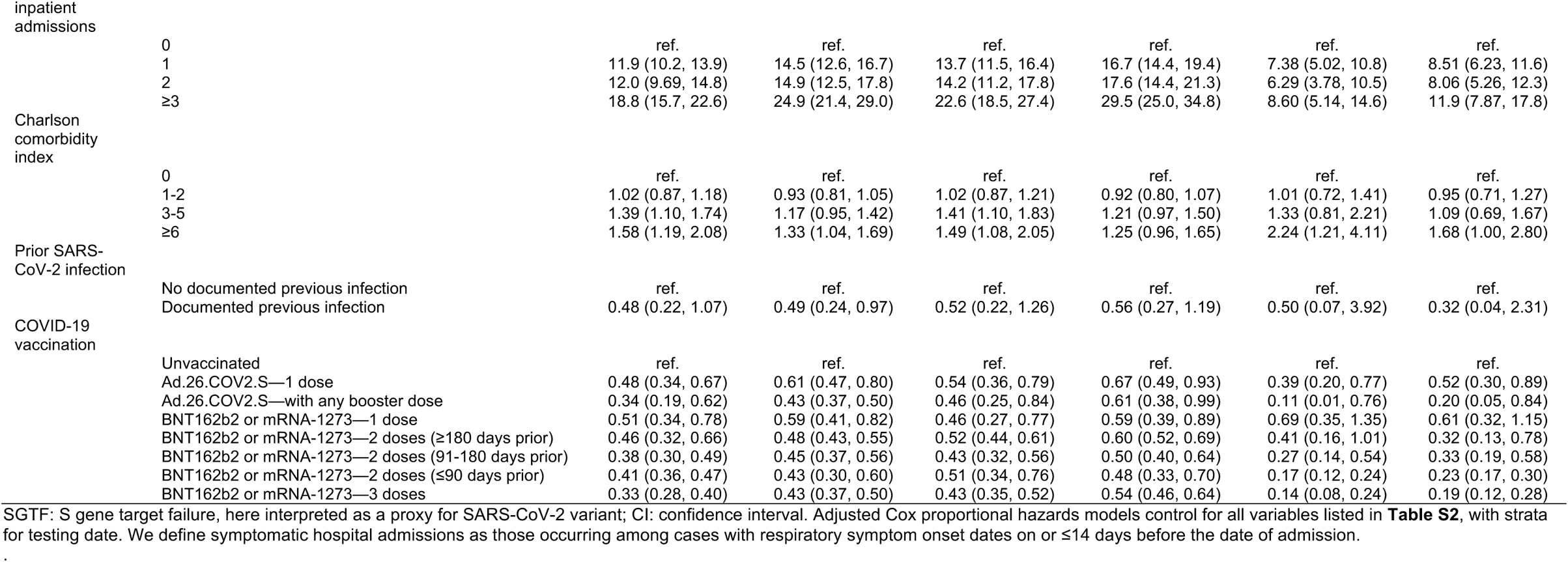
Association of various clinical and demographic characteristics with symptomatic hospital admission among cases with and without SGTF, 15 December, 2021 to 17 January, 2022.

**Table S14:**
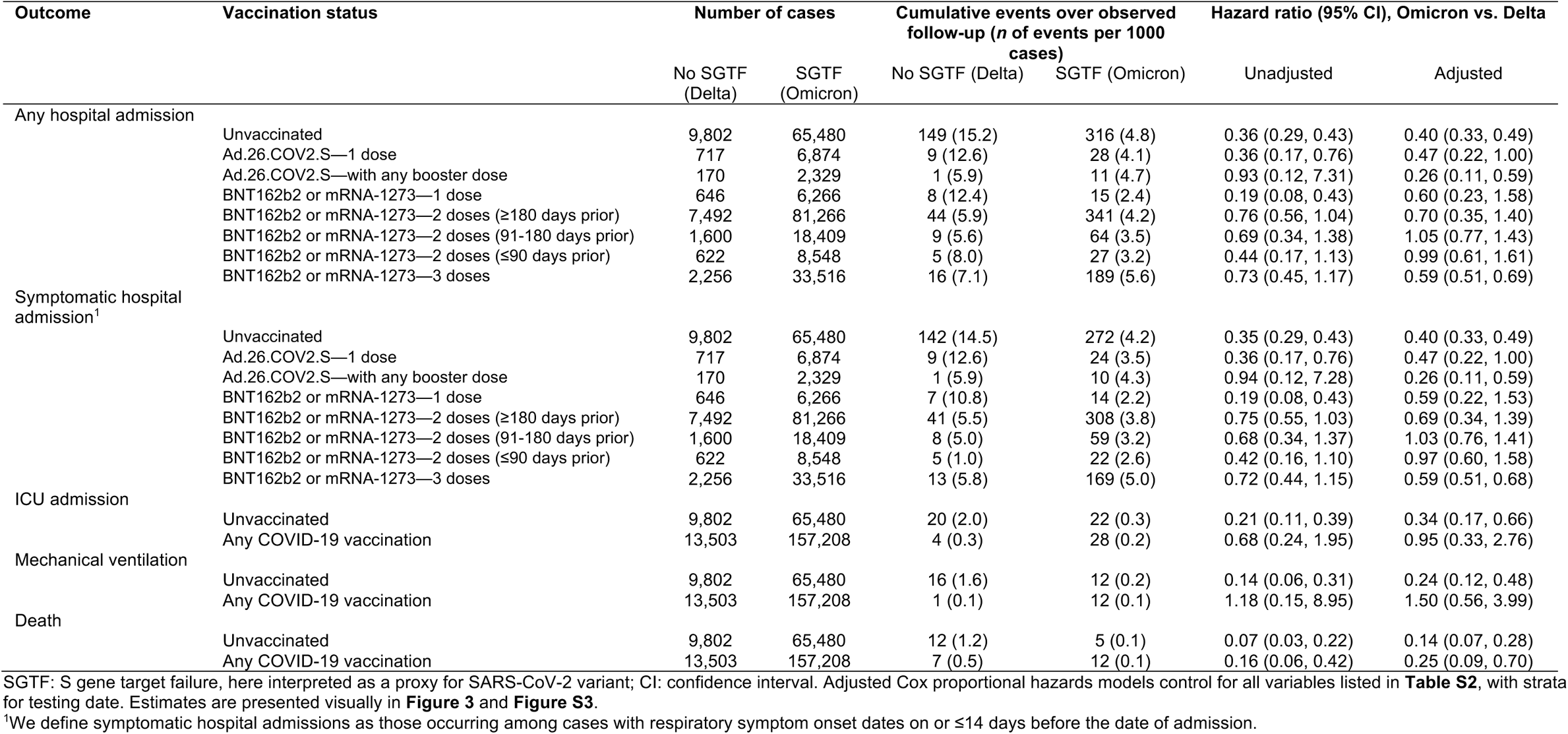
Occurrence of severe clinical outcomes among cases with and without SGTF, stratified by vaccination status, 15 December, 2021 to 17 January, 2022.

**Table S15:** Dates of change in slopes for Cox proportional hazards models relating testing date to risk of severe clinical outcomes.

| Endpoint | Quantile | Changepoint date (95% CI) |
| --- | --- | --- |
| Any hospital admission | Point estimate | 22 December, 2021 |
|  | 25-75%ile | 20 December, 2021 to 24 December, 2021 |
|  | 2.5-97.5%ile | 13 December, 2021 to 3 January, 2022 |
| Symptomatic hospital admission | Point estimate | 23 December, 2021 |
|  | 25-75%ile | 21 December, 2021 to 24 December, 2021 |
|  | 2.5-97.5%ile | 13 December, 2021 to 4 January, 2022 |
| ICU admission | Point estimate | 13 December, 2021 |
|  | 25-75%ile | 7 December, 2021 to 17 December, 2021 |
|  | 2.5-97.5%ile | 19 November, 2021 to 3 January, 2022 |
| Mortality | Point estimate | 8 December, 2021 |
|  | 25-75%ile | 4 December, 2021 to 11 December, 2021 |
|  | 2.5-97.5%ile | 21 November, 2021 to 28 December, 2021 |

CI: Confidence interval. Adjusted Cox proportional hazards models fitted with 0, 1, or 2 changepoints (allowed for each date from 15 November, 2021 to 10 January, 2022) were sampled according to posterior weights. Daily adjusted hazards (controlling for all variables listed in **Table S2**) are illustrated in the inset panels to **Figure 4**.

**Table S16:** Disposition at completion of hospitalization among patients with completed hospital stays.

| Discharge disposition | Number of cases (%) |  |  |  |
| --- | --- | --- | --- | --- |
|  | 15 December, 2021 to 17 January, 2022 |  | 3 February to 17 March, 2022 |  |
|  | No SGTF (Delta) | SGTF (Omicron) | No SGTF (BA.2) | SGTF (BA.1*) |
|  | N=208 | N=703 | N=23 | N=146 |
| Home (self-care) | 138 (66.3) | 582 (82.8) | 20 (87.0) | 124 (84.9) |
| Home with skilled care provision | 22 (10.6) | 36 (5.1) | 2 (8.7) | 8 (5.5) |
| Discharged against medical advice | 4 (1.9) | 1 (0.1) | 0 (0.0) | 2 (1.4) |
| SNF, Residential facility, or rehabilitation facility | 2 (1.0) | 3 (0.4) | 0 (0.0) | 2 (1.4) |
| Other acute inpatient hospital | 2 (1.0) | 2 (0.3) | 0 (0.0) | 1 (0.7) |
| Hospice care/in-hospital death | 2 (1.0) | 3 (0.4) | 0 (0.0) | 0 (0.0) |
| Final disposition not recorded | 38 (18.3) | 76 (10.8) | 1 (4.3) | 9 (6.2) |

SGTF: S gene target failure. Cases in the table are subset to those tested in outpatient settings and admitted within the indicated date ranges, with symptoms onset on or ≤14 days before this date.

**Table S17:** Association of SGTF with time to completion of hospital stay among outpatient-diagnosed cases with symptoms onset on or before admission date, 15 December, 2021 to 17 January, 2022 and 3 February to 17 March, 2022.

| Comparison and period | Hazard ratio (95% CI) |  |
| --- | --- | --- |
|  | Unadjusted | Adjusted |
| Omicron (SGTF) vs. Delta (No SGTF) |  |  |
| 15 December, 2021 to 17 January, 2022 | 1.46 (1.20, 1.77) | 1.24 (0.99, 1.57) |
| BA.2 (No SGTF) vs. BA.1* (SGTF) |  |  |
| 3 February to 17 March, 2022 | 1.02 (0.61, 1.70) | 0.95 (0.41, 2.22) |

SGTF: S gene target failure, here interpreted as a proxy for SARS-CoV-2 variant; CI: confidence interval. Adjusted Cox proportional hazards models control for all variables listed in **Table S2**, with strata for admission date.

**Table S18:**
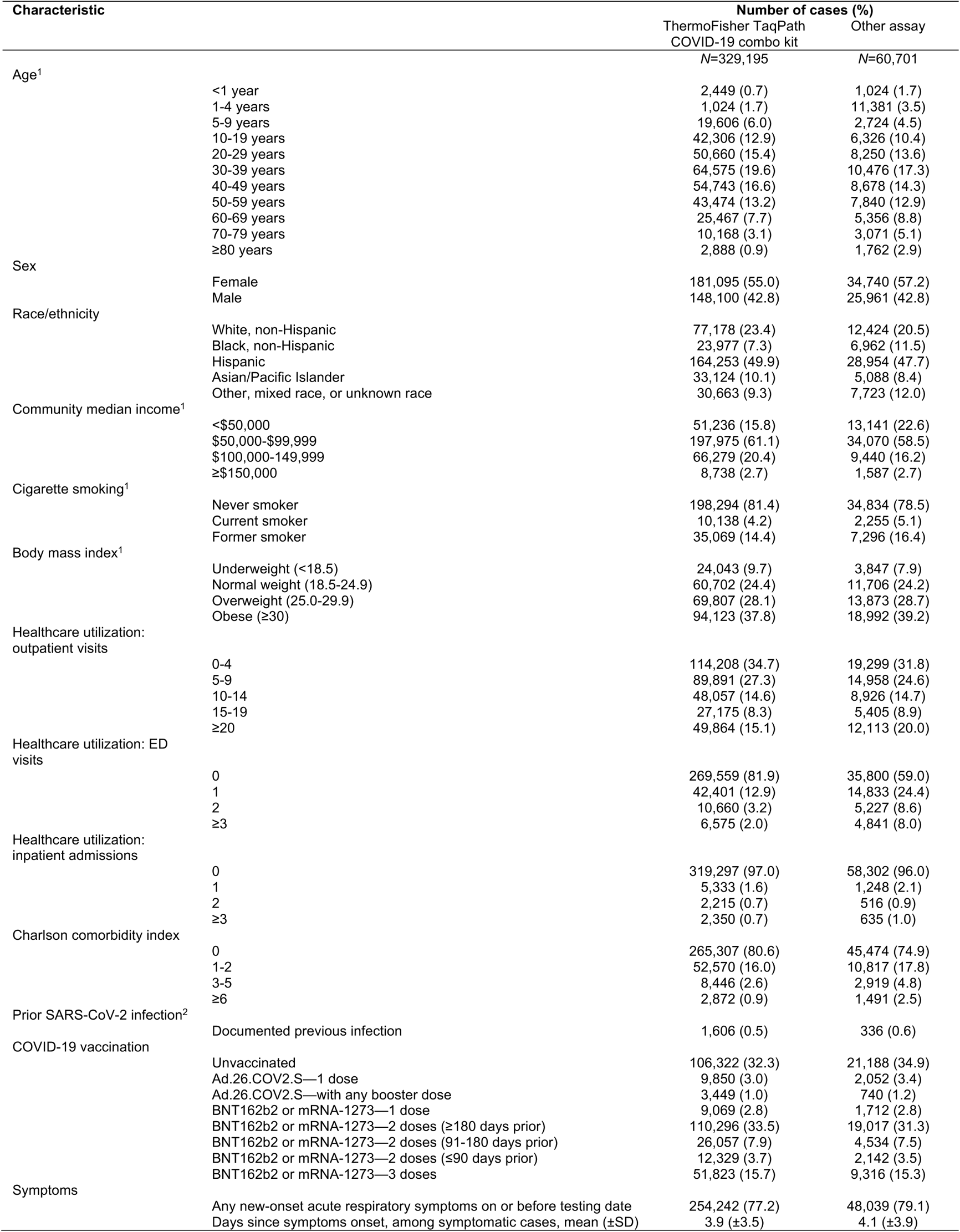
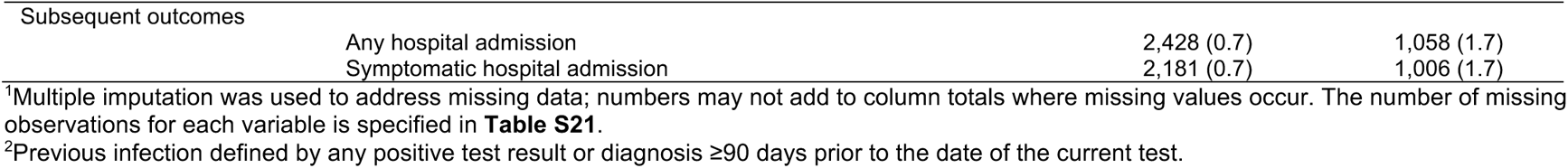
Attributes of outpatient cases with samples processed using the ThermoFisher TaqPath COVID-19 Combo Kit or other assays, 1 November, 2021 to 17 March, 2022.

**Table S19:**
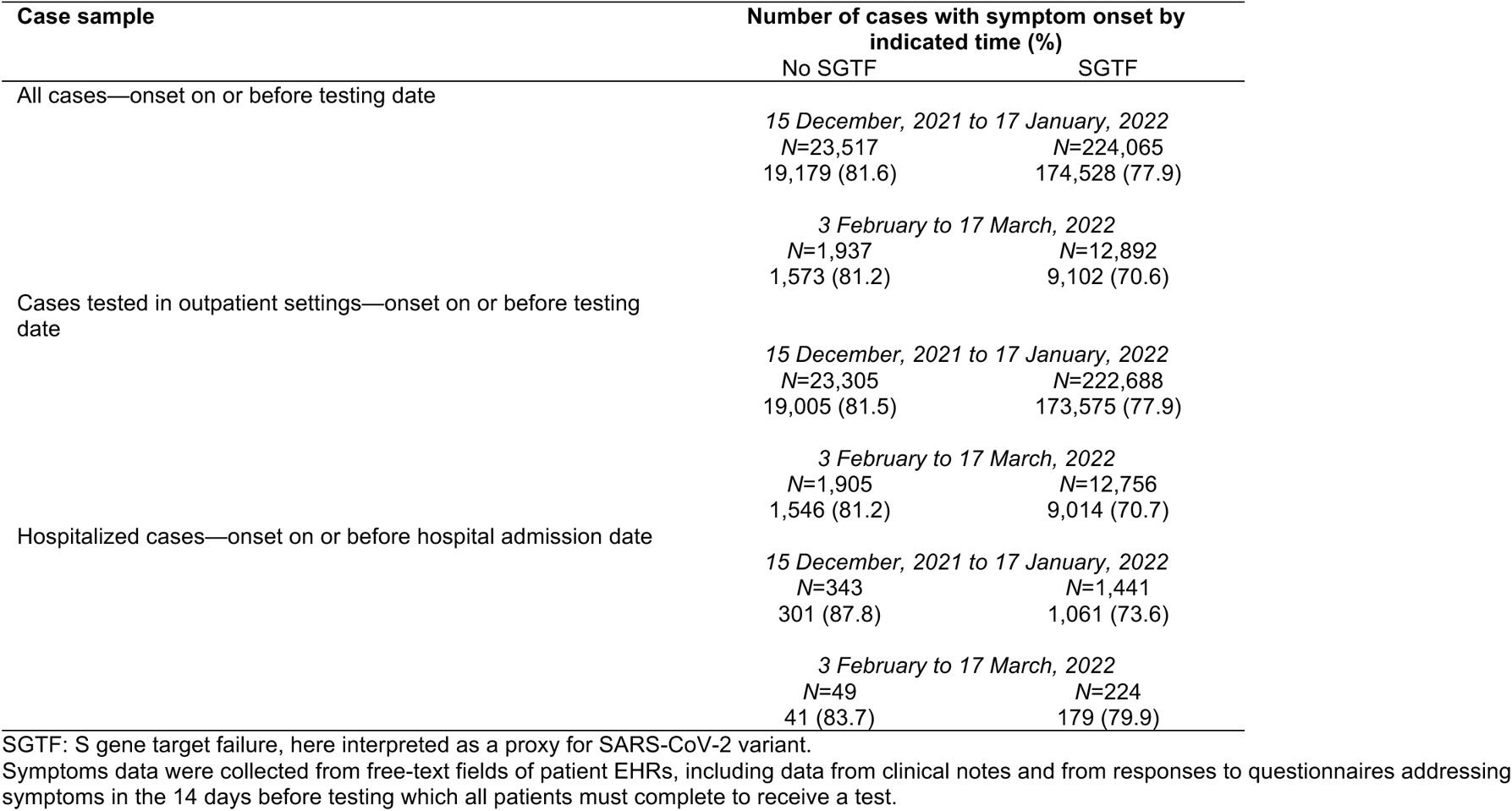
Onset of acute respiratory symptoms on or before dates of testing and hospital admission.

| Case sample | Number of cases with symptom onset by indicated time (%) |  |
| --- | --- | --- |
|  | No SGTF | SGTF |
| All cases—onset on or before testing date | *15 December, 2021 to 17 January, 2022*<br>*N=23,517*<br>19,179 (81.6) | *15 December, 2021 to 17 January, 2022*<br>*N=224,065*<br>174,528 (77.9) |
|  | *3 February to 17 March, 2022*<br>*N=1,937*<br>1,573 (81.2) | *3 February to 17 March, 2022*<br>*N=12,892*<br>9,102 (70.6) |
| Cases tested in outpatient settings—onset on or before testing date | *15 December, 2021 to 17 January, 2022*<br>*N=23,305*<br>19,005 (81.5) | *15 December, 2021 to 17 January, 2022*<br>*N=222,688*<br>173,575 (77.9) |
|  | *3 February to 17 March, 2022*<br>*N=1,905*<br>1,546 (81.2) | *3 February to 17 March, 2022*<br>*N=12,756*<br>9,014 (70.7) |
| Hospitalized cases—onset on or before hospital admission date | *15 December, 2021 to 17 January, 2022*<br>*N=343*<br>301 (87.8) | *15 December, 2021 to 17 January, 2022*<br>*N=1,441*<br>1,061 (73.6) |
|  | *3 February to 17 March, 2022*<br>*N=49*<br>41 (83.7) | *3 February to 17 March, 2022*<br>*N=224*<br>179 (79.9) |

SGTF: S gene target failure, here interpreted as a proxy for SARS-CoV-2 variant.
Symptoms data were collected from free-text fields of patient EHRs, including data from clinical notes and from responses to questionnaires addressing symptoms in the 14 days before testing which all patients must complete to receive a test.

**Table S20:**
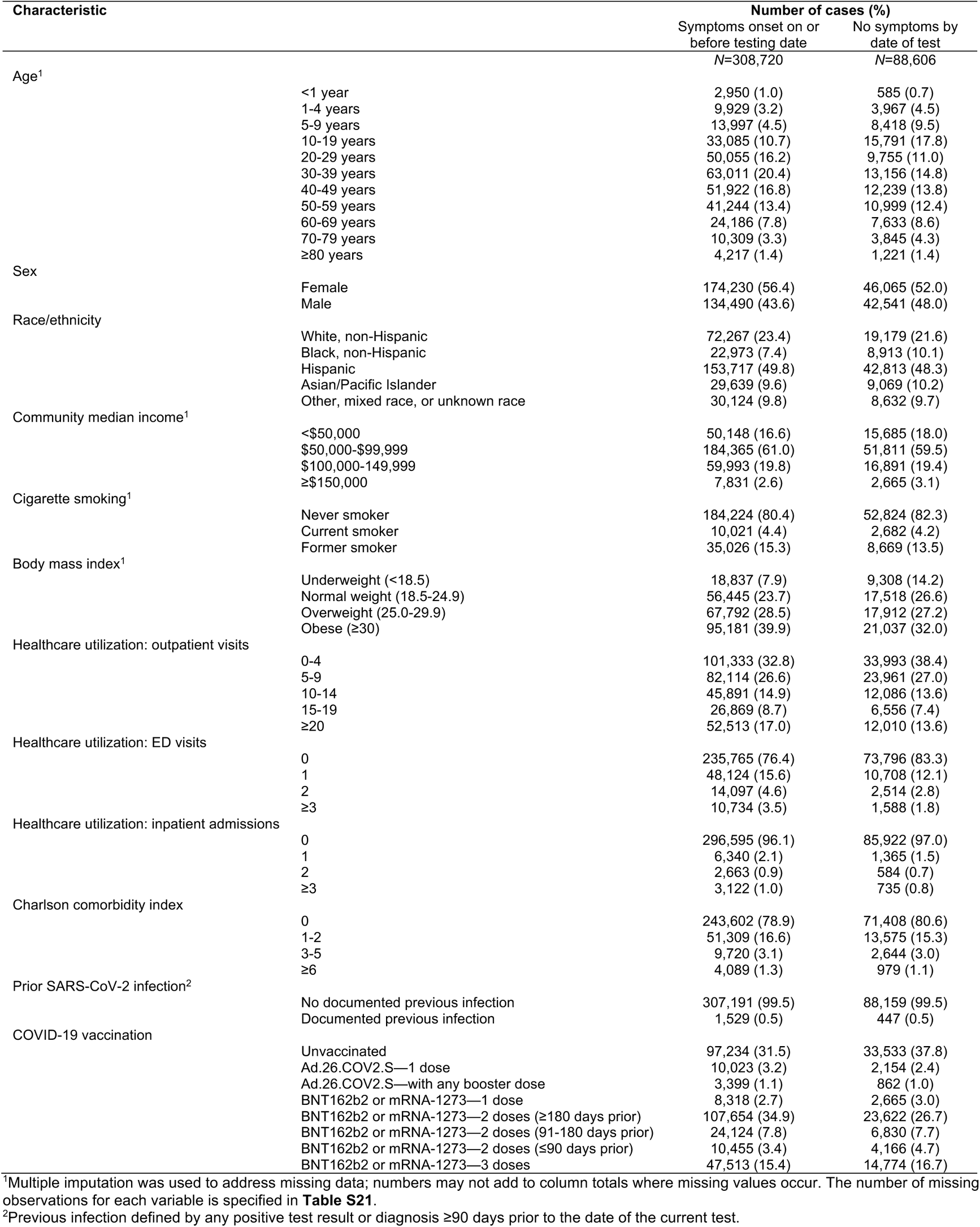
Attributes of outpatient cases with and without symptoms at the time of outpatient testing, 1 November, 2021 to 17 March, 2022.

**Table S21:** Summary of missing data.

| Variable | Missing observations, *n* (%) among cases tested in outpatient settings |  |
| --- | --- | --- |
|  | Full analysis population<sup>1</sup> | Population enrolled ≥1 year |
|  | *N*=329,195 | *N*=283,453 |
| Age | 2,535 (0.8) | 755 (0.3) |
| Sex | 1,057 (0.3) | 755 (0.3) |
| Community median income | 6,024 (1.8) | 1,014 (0.4) |
| History of cigarette smoking | 86,751 (26.3) | 64,961 (22.9) |
| BMI | 81,577 (24.7) | 62,826 (22.1) |
| Any missing observation | 107,870 (32.7) | 81,923 (28.8) |
| Total missing observations, across all covariates analyzed | 188,514 (3.8) | 137,861 (3.2) |

<sup>1</sup>The primary analysis population includes outpatient-tested cases whose samples were processed using the ThermoFisher TaqPath COVID-19 combo kit.

**Table S22:** Association of imputed variables with symptomatic hospital admission endpoint across differing analysis strategies.

| Characteristic | Case population<sup>1</sup> | Primary analysis:<br>Adjusted hazard ratio<br>(95% CI) | Complete case analysis:<br>Adjusted hazard ratio<br>(95% CI) | Participants enrolled<br>≥1 year: Adjusted<br>hazard ratio (95% CI) |
| --- | --- | --- | --- | --- |
| Age | <1 year | 0.12 (0.04, 0.36) | — | — |
|  | 1-4 year | 0.32 (0.16, 0.61) | 0.34 (0.12, 0.94) | 0.28 (0.13, 0.60) |
|  | 5-9 years | 0.27 (0.14, 0.50) | 0.44 (0.23, 0.87) | 0.24 (0.12, 0.48) |
|  | 10-19 years | 0.28 (0.19, 0.41) | 0.31 (0.21, 0.48) | 0.29 (0.20, 0.43) |
|  | 20-29 years | 0.86 (0.71, 1.04) | 0.90 (0.73, 1.10) | 0.84 (0.68, 1.04) |
|  | 30-39 years | ref. | ref. | ref. |
|  | 40-49 years | 0.76 (0.64, 0.92) | 0.70 (0.57, 0.86) | 0.78 (0.64, 0.96) |
|  | 50-59 years | 0.85 (0.70, 1.03) | 0.79 (0.64, 0.98) | 0.90 (0.73, 1.11) |
|  | 60-69 years | 1.34 (1.09, 1.65) | 1.19 (0.95, 1.49) | 1.44 (1.15, 1.79) |
|  | 70-79 years | 1.99 (1.55, 2.56) | 1.79 (1.37, 2.32) | 2.01 (1.55, 2.62) |
|  | ≥80 years | 2.89 (2.08, 3.99) | 2.38 (1.68, 3.39) | 2.88 (2.04, 4.07) |
| Community median income<br>(continuous)<sup>2</sup> | Per 1-log increase | 0.99 (0.85, 1.14) | 1.04 (0.88, 1.23) | 1.01 (0.86, 1.18) |
| Community median income<br>(factorized)<sup>2</sup> | <\$50,000 | 0.95 (0.80, 1.13) | 0.94 (0.79, 1.12) | 0.95 (0.80, 1.13) |
| | \$50,000-\$99,999 | ref. | ref. | ref. |
| | \$100,000-\$149,999 | 0.88 (0.75, 1.04) | 0.91 (0.77, 1.08) | 0.88 (0.74, 1.04) |
| | ≥\$150,000 | 0.64 (0.39, 1.04) | 0.72 (0.45, 1.15) | 0.65 (0.40, 1.05) |
| Cigarette smoking<sup>2</sup> | Never smoker | ref. | ref. | ref. |
|  | Current smoker | 0.97 (0.75, 1.24) | 1.09 (0.82, 1.45) | 0.97 (0.73, 1.30) |
|  | Former smoker | 1.01 (0.87, 1.17) | 1.06 (0.91, 1.24) | 1.04 (0.89, 1.22) |
| Body mass index<sup>2</sup> | Underweight (<18.5) | 0.80 (0.51, 1.27) | 0.72 (0.39, 1.33) | 0.91 (0.56, 1.49) |
|  | Normal weight (18.5-24.9) | ref. | ref. | ref. |
|  | Overweight (25.0-29.9) | 1.35 (1.13, 1.62) | 1.47 (1.21, 1.79) | 1.33 (1.10, 1.61) |
|  | Obese (≥30) | 1.60 (1.36, 1.90) | 1.71 (1.42, 2.07) | 1.61 (1.34, 1.94) |

CI: confidence interval. Adjusted Cox proportional hazards models control for all variables listed in **Table S2**, with strata for testing date
<sup>1</sup>The primary analysis population includes outpatient-tested cases whose samples were processed using the ThermoFisher TaqPath COVID-19 combo kit.
<sup>2</sup>Primary analyses addressed community median household income as a continuous variable; we present the factorized version (broken down by increments of \$50,000/year) to further illustrate similarity of effect estimates across the analysis strategies.

**Figure S1:**
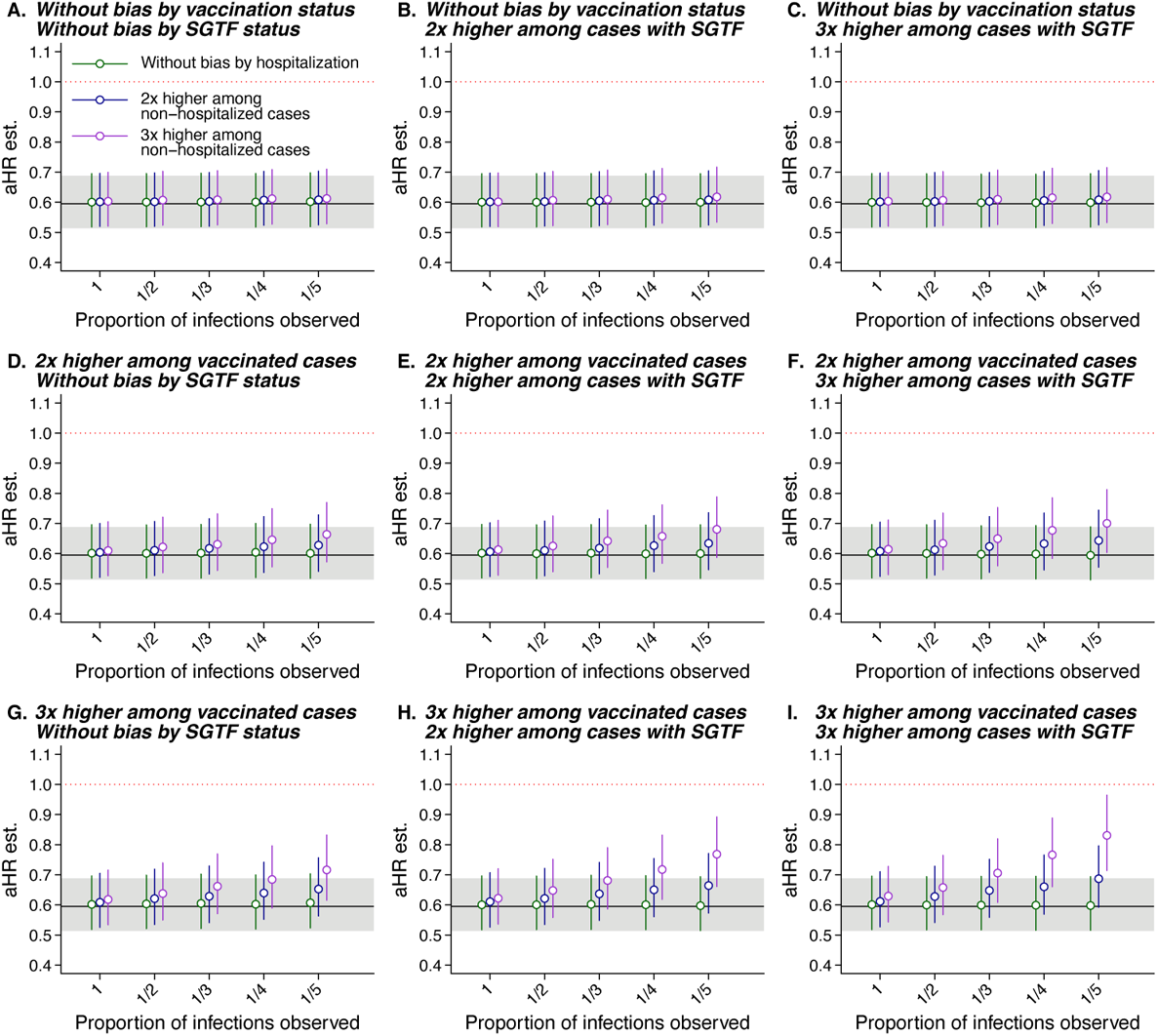
Estimates of the adjusted hazard ratio (aHR) of any hospital admission under scenarios of differential unobserved prior infection prevalence across case strata. We illustrate values of the aHR of any hospital admission among outpatient diagnosed cases with Omicron vs. Delta infection, estimated in analyses allowing for differing levels of prior infection prevalence according to their infecting variant, outcome (hospital admission or no hospital admission), and history of vaccination. Panels indicate the fold elevation in infection prevalence applied to each stratum over a baseline proportion of infections assumed to be unascertained on the x-axis for unvaccinated cases with Delta variant infection who were hospitalized (highest-risk stratum). Horizontal lines at aHR=0.59 and accompanying gray shaded areas delineate the point estimate and 95% confidence interval from primary analysis models.

**Figure S2:**
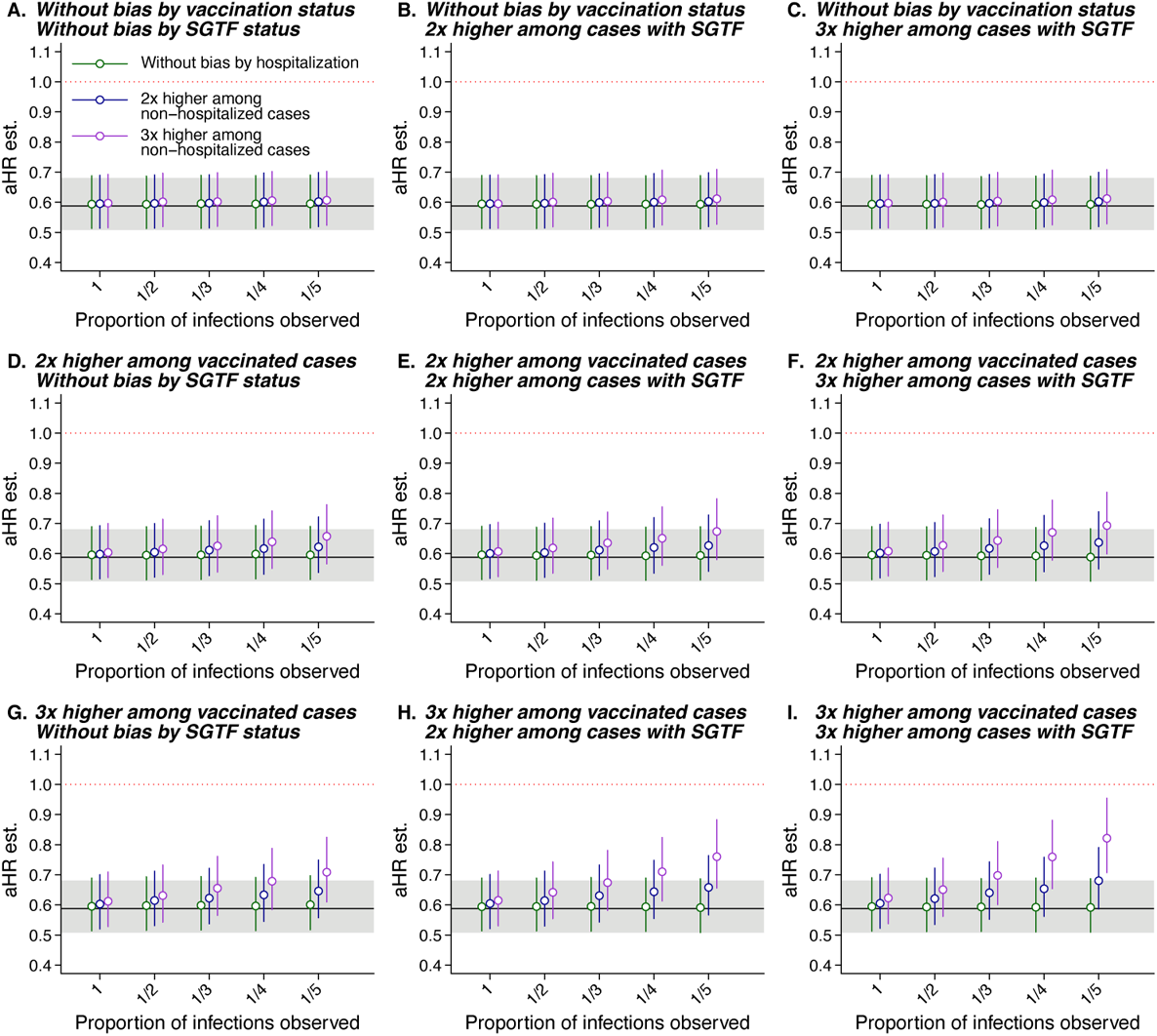
Estimates of the adjusted hazard ratio (aHR) of symptomatic hospital admission under scenarios of differential unobserved prior infection prevalence across case strata. We illustrate values of the aHR of any hospital admission among outpatient diagnosed cases with Omicron vs. Delta infection, estimated in analyses allowing for differing levels of prior infection prevalence according to their infecting variant, outcome (symptomatic hospital admission or no symptomatic hospital admission), and history of vaccination. Panels indicate the fold elevation in infection prevalence applied to each stratum over a baseline proportion of infections assumed to be unascertained on the x-axis for unvaccinated cases with Delta variant infection who were hospitalized (highest-risk stratum). Horizontal lines at aHR=0.59 and accompanying gray shaded areas delineate the point estimate and 95% confidence interval from primary analysis models.

**Figure S3:**
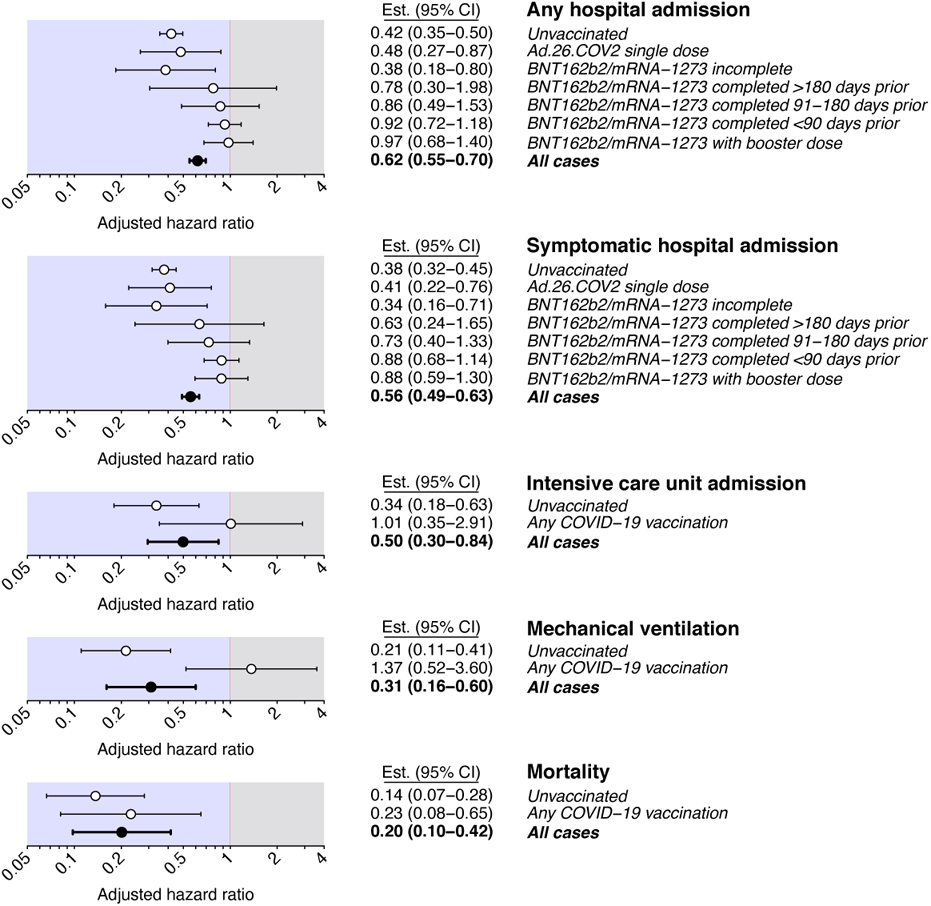
Adjusted hazard ratios of severe clinical endpoints associated with Omicron variant infection, without restriction by diagnosis setting. Points and lines denote estimates and accompanying 95% confidence intervals for the adjusted hazard ratio of each endpoint, comparing cases with Omicron versus Delta variant infection, in case strata defined by history of COVID-19 vaccination. Analyses are restricted to individuals tested diagnosed by RT-PCR testing using the ThermoFisher TaqPath COVID-19 combo kit, in contrast to **Figure 3** which further limits analyses to cases diagnosed in outpatient settings. Adjusted hazard ratios are estimated using Cox proportional hazards regression models, controlling for covariates listed in Table S2 and defining strata for testing date.

**Figure S4:**
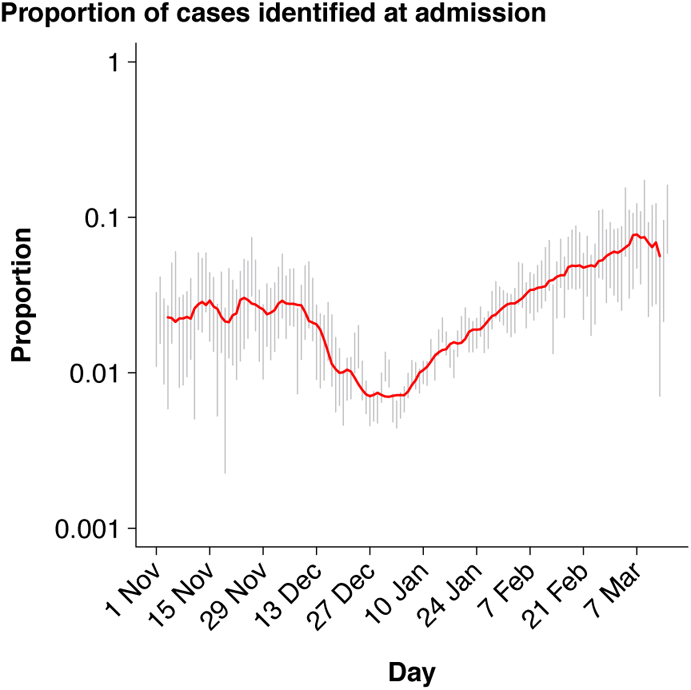
Changes in the proportion of cases ascertained on or after the date of hospital admission. We illustrate day-specific estimates of the proportion of cases ascertained on or after the hospital admission date with 95% confidence intervals generated by bootstrap sampling (grey lines) as well as the 7-day moving average (red line).

## Notes

### Competing Interest Statement

JAL discloses receipt of grants from Pfizer and Merck, Sharp & Dohme, and consulting fees from Pfizer, Merck, Sharp & Dohme, and VaxCyte, Inc., unrelated to this work. SYT discloses receipt of grants from Pfizer unrelated to this work. ML reports grants from CDC, grants from NIH, grants from UK NIHR, grants from Pfizer, personal fees from Merck, personal fees from Janssen, personal fees from Sanofi Pasteur, personal fees from Bristol Myers Squibb, personal fees from Peter Diamandis/Abundance Platinum, outside the submitted work; and Unpaid advice to One Day Sooner, Pfizer, Janssen, Astra-Zeneca, Covaxx (United Biomedical). VXH, MMP, and RK have no competing interests to disclose.

### Funding Statement

The study was funded by the US Centers for Disease Control and Prevention.

### Author Declarations

The study protocol was reviewed and approved by the Kaiser Permanente Southern California Institutional Review Board, which waived requirement for informed consent.

## References

1. World Health Organization. Update on Omicron variant. (2021). Accessed 10 January, 2022. https://www.who.int/news/item/28-11-2021-update-on-omicron

2. Karim, S. S. A. & Karim, Q. A. Omicron SARS-CoV-2 variant: a new chapter in the COVID-19 pandemic. Lancet 398, 2126–2128 (2021).

3. Pearson, C. A. B. et al. Bounding the levels of transmissibility and immune evasion of the Omicron variant in South Africa. medRxiv (2021). doi:10.1101/2021.12.19.21268038.

4. Iuliano, A. D. et al. Trends in disease severity and health care utilization during the early omicron variant period compared with previous SARS-CoV-2 high transmission periods—United States, December 2020--January 2022. Morbid Mortal Wkly Rep (2022). doi:10.15585/mmwr.mm7104e4.

5. Public Health England. SARS-CoV-2 variants of concern and variants under investigation in England—Technical briefing: Update on hospitalisation and vaccine effectiveness for Omicron VOC-21NOV-01 (B.1.1.529). (2021). Accessed 21 February, 2022, https://assets.publishing.service.gov.uk/government/uploads/system/uploads/attachment_data/file/1045619/Technical-Briefing-31-Dec-2021-Omicron_severity_update.pdf.

6. Madhi, S. A. et al. Population immunity and COVID-19 severity with Omicron variant in South Africa. NEJM (2022). doi:10.1056/nejmoa2119658.

7. Bhattacharyya, R. P. & Hanage, W. P. Challenges in inferring intrinsic severity of the SARS-CoV-2 Omicron variant. NEJM (2022). doi:10.1056/NEJMp2119682.

8. First confirmed case of Omicron variant detected in the United States. US Centers for Disease Control and Prevention (2021). Accessed 10 January, 2022. https://www.cdc.gov/media/releases/2021/s1201-omicron-variant.htm

9. US Centers for Disease Control and Prevention. COVID-19 Data Tracker: Variant proportions (2022). Accessed 8 May, 2022. https://covid.cdc.gov/covid-data-tracker/#variant-proportions.

10. Taylor, L. Covid-19: Omicron drives weekly record high in global infections. BMJ (2022). doi:10.1136/bmj.o66.

11. Clarke, K. E. N. et al. Seroprevalence of infection-induced SARS-CoV-2 antibodies — United States, September, 2021–February, 2022. Morbid Mortal Wkly Rep (2022). doi:10.15585/mmwr.mm7117e3.

12. US Centers for Disease Control and Prevention. COVID-19 Data Tracker: Trends in number of COVID-19 cases and deaths in the US reported to CDC, by state/territory (2022). Accessed 8 May, 2022. https://covid.cdc.gov/covid-data-tracker/#trends_dailycases

13. Matthew, M. et al. Structural basis of SARS-CoV-2 Omicron immune evasion and receptor engagement. Science 375, 864–868 (2022).

14. Nemet, I. et al. Third BNT162b2 vaccination neutralization of SARS-CoV-2 Omicron infection. N Engl J Med (2021). doi:10.1056/NEJMc2119358.

15. Schmidt, F. et al. Plasma neutralization of the SARS-CoV-2 Omicron variant. N Engl J Med (2021). doi:10.1056/NEJMc2119641.

16. Altarawneh, H. N. et al. Protection against the Omicron variant from previous SARS-CoV-2 infection. N Engl J Med (2022). doi:10.1056/NEJMc2200133.

17. Pulliam, J. R. C. et al. Increased risk of SARS-CoV-2 reinfection associated with emergence of Omicron in South Africa. Science (2022). doi:10.1126/science.abn4947.

18. Tseng, H. F. et al. Effectiveness of mRNA-1273 against SARS-CoV-2 Omicron and Delta variants. Nat Med (2022). doi:10.1038/s41591-022-01753-y.

19. Tartof, S. Y. et al. Durability of BNT162b2 vaccine against hospital and emergency department admissions due to the Omicron and Delta variants in a large health system in the USA: a test-negative case-control study. Lancet Respir Med (2022). doi:10.1016/S2213-2600(22)00101-1.

20. Nyberg, T. et al. Comparative analysis of the risks of hospitalisation and death associated with SARS-CoV-2 Omicron (B.1.1.529) and Delta (B.1.617.2) variants in England: a cohort study. Lancet 399, 1303–1312 (2022).

21. Bager, P. et al. Reduced risk of hospitalisation associated with infection with SARS-CoV-2 Omicron variant versus Delta variant in Denmark: an observational cohort study. Lancert Infect Dis (2022). doi:10.1016/S1473-3099(22)00154-2.

22. Wolter, N. et al. Early assessment of the clinical severity of the SARS-CoV-2 omicron variant in South Africa: a data linkage study. Lancet 399, 437–446 (2022).

23. Qassim, S. H. et al. Effects of BA.1/BA.2 subvariant, vaccination, and prior infection on infectiousness of SARS-CoV-2 Omicron infections. medRxiv (2022). doi:10.1101/2022.03.02.22271771.

24. Wolter, N., Jassat, W., author group, D.-G., von Gottberg, A. & Cohen, C. Clinical severity of Omicron sub-lineage BA.2 compared to BA.1 in South Africa. medRxiv (2022). doi:10.1101/2022.02.17.22271030.

25. Lesley, S. et al. Track Omicron’s spread with molecular data. Science 374, 1454–1455 (2021).

26. Andrews, N. et al. Covid-19 vaccine effectiveness against the Omicron (B.1.1.529) variant. NEJM 386, 1532–1546 (2022).

27. Stensrud, M. J. & Hernán, M. A. Why test for proportional hazards? JAMA 323, 1401–1402 (2020).

28. Kürüm, E. et al. Bayesian model averaging with change points to assess the impact of vaccination and public health interventions. Epidemiology 28, 889–897 (2017).

29. Strålin, K. et al. Impact of the Alpha VOC on disease severity in SARS-CoV-2-positive adults in Sweden. J Infect 84, e3–e5 (2022).

30. Garvey, M. I. et al. Observations of SARS-CoV-2 variant of concern B.1.1.7 at the UK’s largest hospital trust. J Infect 83, e21–e23 (2021).

31. Sheikh, A. et al. Severity of Omicron variant of concern and effectiveness of vaccine boosters against symptomatic disease in Scotland (EAVE II): a national cohort study with nested test-negative design. Lancet Infect Dis (2022). doi:10.1016/S1473-3099(22)00141-4.

32. Lauring, A. S. et al. Clinical severity of, and effectiveness of mRNA vaccines against, Covid-19 from Omicron, Delta, and Alpha SARS-CoV-2 variants in the United States: prospective observational study. BMJ 376, (2022).

33. Ulloa, A. C., Buchan, S. A., Daneman, N. & Brown, K. A. Estimates of SARS-CoV-2 Omicron Variant Severity in Ontario, Canada. JAMA 327, 1286–1288 (2022).

34. Peralta-Santos, A. et al. Omicron (BA.1) SARS-CoV-2 variant is associated with reduced risk of hospitalization and length of stay compared with Delta (B.1.617.2). medRxiv (2022). doi:10.1101/2022.01.20.22269406.

35. Laxminarayan, R. et al. SARS-CoV-2 infection and mortality during the first epidemic wave in Madurai, south India: a prospective, active surveillance study. Lancet Infect Dis 21, 1665–1676 (2021).

36. Davies, N. G. et al. Increased mortality in community-tested cases of SARS-CoV-2 lineage B.1.1.7. Nature 593, 270–274 (2021).

37. Davies, N. G. et al. Estimated transmissibility and impact of SARS-CoV-2 lineage B.1.1.7 in England. Science 372, eabg3055 (2021).

38. Huynh, D. N. et al. Description and early results of the Kaiser Permanente Southern California COVID-19 home monitoring program. Perm. J. 25, 1–7 (2021).

39. Klann, J. G. et al. Distinguishing admissions specifically for COVID-19 from incidental SARS-CoV-2 admissions: a national retrospective EHR study. medRxiv (2022). doi:10.1101/2022.02.10.22270728.

40. Nyberg, T. et al. Risk of hospital admission for patients with SARS-CoV-2 variant B.1.1.7: cohort analysis. BMJ 373, n1412 (2021).

41. Sigal, A., Milo, R. & Jassat, W. Estimating disease severity of Omicron and Delta SARS-CoV-2 infections. Nat Rev Immunol (2022). doi:10.1038/s41577-022-00720-5.

42. Bhattacharyya, R. P. & Hanage, W. P. Challenges in inferring intrinsic severity of the SARS-CoV-2 Omicron variant. NEJM (2022). doi:10.1056/NEJMp2119682.

43. Lamba, K., et al. SARS-CoV-2 cumulative incidence and period seroprevalence: results from a statewide population-based serosurvey in California. Open Forum Infect. Dis. 8, ofab379 (2021).

44. Carreño, J. M. et al. Activity of convalescent and vaccine serum against SARS-CoV-2 Omicron. Nature (2022). doi:10.1038/s41586-022-04399-5.

45. Rössler, A., Riepler, L., Bante, D., von Laer, D. & Kimpel, J. SARS-CoV-2 Omicron variant neutralization in serum from vaccinated and convalescent persons. N Engl J Med (2022). doi:10.1056/nejmc2119236.

46. Griffith, G. J. et al. Collider bias undermines our understanding of COVID-19 disease risk and severity. Nat Commun 11, 5749 (2020).

47. Vandenbroucke, J. P., Broadbent, A. & Pearce, N. Causality and causal inference in epidemiology: The need for a pluralistic approach. Int J Epidemiol 45, 1776–1786 (2016).

48. Hui, K. P. Y. et al. SARS-CoV-2 Omicron variant replication in human bronchus and lung ex vivo. Nature (2022). doi:10.1038/s41586-022-04479-6.

49. Abdelnabi, R. et al. The Omicron (B.1.1.529) SARS-CoV-2 variant of concern does not readily infect Syrian hamsters. Antiviral Res 198, 105253 (2022).

50. Gozzi, N. et al. Preliminary modeling estimates of the relative transmissibility and immune escape of the Omicron SARS-CoV-2 variant of concern in South Africa. medRxiv (2022). doi:10.1101/2022.01.04.22268721.

51. Smith, D. J. et al. COVID-19 mortality and vaccine coverage — Hong Kong. Morbid Mortal Wkly Rep 71, 545–548 (2022).

52. Statistics New Zealand. COVID-19 data portal. Accessed 8 May 2022, https://www.stats.govt.nz/experimental/covid-19-data-portal.

53. The COVID Tracking Project. Long-term-care COVID tracker. Accessed 8 May 2022, https://covidtracking.com/nursing-homes-long-term-care-facilities.

## Additional references

54. Tartof, S. Y. et al. Effectiveness of mRNA BNT162b2 COVID-19 vaccine up to 6 months in a large integrated health system in the USA: a retrospective cohort study. Lancet 398, 1407–1416 (2021).

55. Koebnick, C. et al. Sociodemographic characteristics of members of a large, integrated health care system: comparison with US Census Bureau data. Perm J 16, 37–41 (2012).

56. California Department of Public Health. Variants in California — COVID-19 Response. Accessed 8 May 2022, https://covid19.ca.gov/variants/.

57. Rubin, D. B. Multiple imputation after 18+ years. J Am Stat Assoc 91, 473–489 (1996).

58. León, T. M. et al. COVID-19 cases and hospitalizations by COVID-19 vaccination status andprevious COVID-19 diagnosis — California and New York, May–November 2021. Morbid Mortal Wkly Rep 71, 125–131 (2022).

59. Andrejko, K. L. et al. Prevention of COVID-19 by mRNA-based vaccines within the general population of California. Clin Infect Dis (2021). doi:10.1093/cid/ciab640

60. Feikin, D. R. Duration of effectiveness of vaccines against SARS-CoV-2 infection and COVID-19 disease: results of a systematic review and meta-regression. Lancet 399, 924–944 (2022).

61. Burnham, K. P. & Anderson, D. R. Multimodel inference: understanding AIC and BIC in model selection. Sociol. Methods Res 33, 261–304 (2004).

62. Therneau, T. M. survival: A Package for Survival Analysis in R. Version 3.3-1 (2022).

63. Honaker, J., King, G. & Blackwell, M. Amelia II: A program for missing data. J Stat Softw (2011). doi:10.18637/jss.v045.i07.

